# Surveillance of SARS-CoV-2 variants in Argentina: detection of Alpha, Gamma, Lambda, Epsilon and Zeta in locally transmitted and imported cases

**DOI:** 10.1101/2021.07.19.21260779

**Authors:** Torres Carolina, Mojsiejczuk Laura, Acuña Dolores, Alexay Sofía, Amadio Ariel, Aulicino Paula, Debat Humberto, Fernández Franco, Goya Stephanie, König Guido, Nabaes Jodar Mercedes, Pianciola Luis, Bengoa Sofía, Cacciahue Marco, Camussone Cecilia, Dus Santos María José, Eberhardt María Florencia, Fernandez Ailen, Gismondi María Inés, Irazoqui Matías, Lusso Silvina, Marquez Nathalie, Muñoz Marianne, Natale Mónica, Pisano Belén, Puebla Andrea, Re Viviana, Sosa Ezequiel, Zaiat Jonathan, Zunino Sebastián, Do porto Darío, Acevedo María Elina, Alvarez Lopez Cristina, Álvarez María Laura, Angeleri Patricia, Angelletti Andrés, Arca Manuel, Barbas Gabriela, Bertone Ana, Bonnet Agustina, Bourlot Ignacio, Castello Alejandro, Castro Gonzalo, Ceriani Carolina, Cimino Carlos, Cipelli Julián, Colmeiro María, Cordero Andrés, Cristina Carolina, Di Bella Sofia, Ercole Regina, Espasandin Yesica, Espul Carlos, Falaschi Andrea, Fernandez Moll Facundo, Gatelli Andrea, Goñi Sandra, Jofré María Estela, Jaramillo José, Labarta Natalia, Lacaze María Agustina, Larreche Rocio, Leiva Viviana, Levin Gustavo, Luczak Erica, Mandile Marcelo, Massone Carla, Mazzeo Melina, Medina Carla, Monaco Belén, Montoto Luciana, Mugna Viviana, Musto Alejandra, Ojeda Guillermo, Pintos Carolina, Pozzati Marcia, Rahhal Marilina, Rechimont Claudia, Remes Lenicov Federico, Rompato Gabriela, Seery Vanesa, Siri Leticia, Spina Julieta, Streitenberger Cintia, Suárez Ariel, Suárez Jorgelina, Sujanski Paula, Talia Juan Manuel, Theaux Clara, Thomas Guillermo, Ticeira Marina, Tittarelli Estefanía, Toro Rosana, Uez Osvaldo, Zaffanella María Belén, Ziehm Cecilia, Zubieta Martin, on behalf of PAIS Consortium, Mistchenko Alicia, Valinotto Laura, Viegas Mariana

## Abstract

Molecular surveillance of SARS-CoV-2 variants was performed on a total of 2,406 samples from the capital city and nine provinces of Argentina, during 30 epidemiological weeks (EW) that covered the end of the first wave and the beginning of the ongoing second wave of the COVID-19 pandemic in the country (EW 44/2020 to EW 20/2021). The surveillance strategy was mainly based on Sanger sequencing of a Spike coding region that allows the simultaneous identification of signature mutations associated with worldwide circulating variants. In addition, whole SARS-CoV-2 genome sequences were obtained from 456 samples. The main variants found were Gamma, Lambda and Alpha, and to a lesser extent, Zeta and Epsilon. Whereas Gamma dominated in different regions of the country, both Gamma and Lambda prevailed in the most populated area, the metropolitan region of Buenos Aires (MABA), although showing a heterogeneous distribution along this region. This cost-effective surveillance protocol allowed for a rapid response in a limited access to resources scenario, added information on the expansion of the Lambda variant in South America and contributed to the implementation of public health measures to control the disease spread in Argentina.

## Main text

The emergence of SARS-CoV-2 variants with concerning characteristics to public health has attracted the attention of the scientific community and governments both regionally and globally since the end of 2020. The most relevant variants described so far include: Alpha (lineage B.1.1.7), first detected in the United Kingdom; Beta (lineage B.1.351), initially detected in South Africa; Gamma (lineage P.1, derived from lineage B.1.1.28), initially detected in Manaus, Brazil, and Japan; Delta (lineage B.1.627.2), initially detected in India; Epsilon (lineages B.1.427 and B.1.429), initially detected in California, United States; Zeta (lineage P.2, derived from lineage B.1.1.28), first detected in Rio de Janeiro, Brazil; and Lambda (lineage C.37, derived from B.1.1.1), initially detected in Peru (1). Four of these variants (Alpha to Delta) have been defined as variants of concern (VOC) given their increased transmissibility and other characteristics (1). They have also been associated with an increased risk of hospitalization (2,3) and, in the case of Beta, Gamma and Delta, with a moderate to substantial reduction in neutralizing activity of monoclonal antibodies, convalescent and vaccine sera (4–6).

Gamma and Lambda are particularly relevant for Argentinean public health due to their significant presence in the South American region. Importantly, some of these variants share distinct mutations in the Spike protein -several of them in the receptor binding domain (RBD) region-that potentially affect transmissibility, pathogenesis and/or response to vaccination and immune-based therapies.

PAIS is the inter-institutional federal consortium of SARS-CoV-2 genomics in Argentina. It was created by the Ministry of Science and Technology to monitor SARS-CoV-2 diversity and evolution in the country, including surveillance of SARS-CoV-2 variants of public health interest (http://pais.qb.fcen.uba.ar/).

Between October 26^th^, 2020 and May 22^nd^, 2021 (epidemiological week (EW)44/2020 to EW20/2021), molecular surveillance was performed on a total of 2,406 samples from the capital city and nine provinces of the country, including the four most populated districts (Figure 1 and Table 1). This period covers the end of the first wave and the beginning of the ongoing second and largest wave of the COVID-19 pandemic in Argentina (Figure 2). During that period, the frontiers were mostly open for Argentinean residents, but the foreigners had severe restrictions to enter the country as tourists.

**Table 1.** Cumulative cases analyzed in this work from different regions of Argentina.

| Region | Area or city | Period | Alpha | Gamma | Lambda | E484K | L452R | Non<br>VOC/VOI | TOTAL |
| --- | --- | --- | --- | --- | --- | --- | --- | --- | --- |
| **CABA (part of MABA)** |  | 2020-10-26 to 2021-05-22 | 81 | 121 | 160 | 67 | 10 | 427 | 866 |
| **GBA (part of MABA)** | North | 2020-12-21 to 2021-05-21 | 5 | 16 | 10 | 4 | 0 | 32 | 67 |
|  | West | 2020-11-28 to 2021-05-22 | 35 | 50 | 59 | 17 | 3 | 108 | 272 |
|  | South | 2020-10-26 to 2021-05-22 | 10 | 47 | 138 | 27 | 5 | 163 | 390 |
| **GLP (part of MABA)** |  | 2020-11-27 to 2021-04-24 | 1 | 43 | 13 | 1 | 2 | 31 | 91 |
| **MABA** | unknown | 2021-02-14 to 2021-05-21 | 3 | 2 | 7 | 0 | 0 | 7 | 19 |
| **PBA (outside MABA)** | Several cities | 2020-11-27 to 2021-05-21 | 42 | 44 | 48 | 17 | 3 | 82 | **236** |
| **Córdoba** | Several cities | 2020-11-03 to 2021-05-21 | 18 | 23 | 16 | 4 | 11 | 83 | **155** |
| **Santa Fe** | Several cities | 2020-12-11 to 2021-05-22 | 9 | 68 | 6 | 0 | 3 | 59 | **145** |
| **Entre Ríos** | Several cities | 2021-05-11 to 2021-05-14 | 11 | 20 | 3 | 0 | 0 | 3 | **37** |
| **La Pampa** | Santa Rosa | 2021-05-16 to 2021-05-17 | 1 | 11 | 1 | 0 | 0 | 0 | **13** |
| **San Luis** | Several cities | 2021-04-02 to 2021-04-17 | 3 | 14 | 3 | 0 | 0 | 10 | **30** |
| **Mendoza** | General Alvear | 2021-03-22 to 2021-04-08 | 0 | 15 | 0 | 0 | 0 | 0 | **15** |
| **Neuquén** | Several cities | 2021-01-03 to 2021-05-15 | 1 | 25 | 1 | 0 | 2 | 33 | **62** |
| **Río Negro** | Bariloche | 2021-12-21 to 2021-03-09 | 0 | 0 | 0 | 1 | 0 | 7 | **8** |
| **TOTAL** |  | **2020-10-26 to 2021-05-22** | **220** | **499** | **465** | **138** | **39** | **1045** | **2406** |

Note: The total in CABA (n = 866) does not include two co-infections (Alpha/Lambda) and the total in GLP (n = 91) does not include one co-infection (Alpha/Gamma).
CABA: Buenos Aires city. GBA: Great Buenos Aires. GLP: Great La Plata. MABA: Metropolitan area of Buenos Aires. PBA: Buenos Aires province.

**Figure 1.**
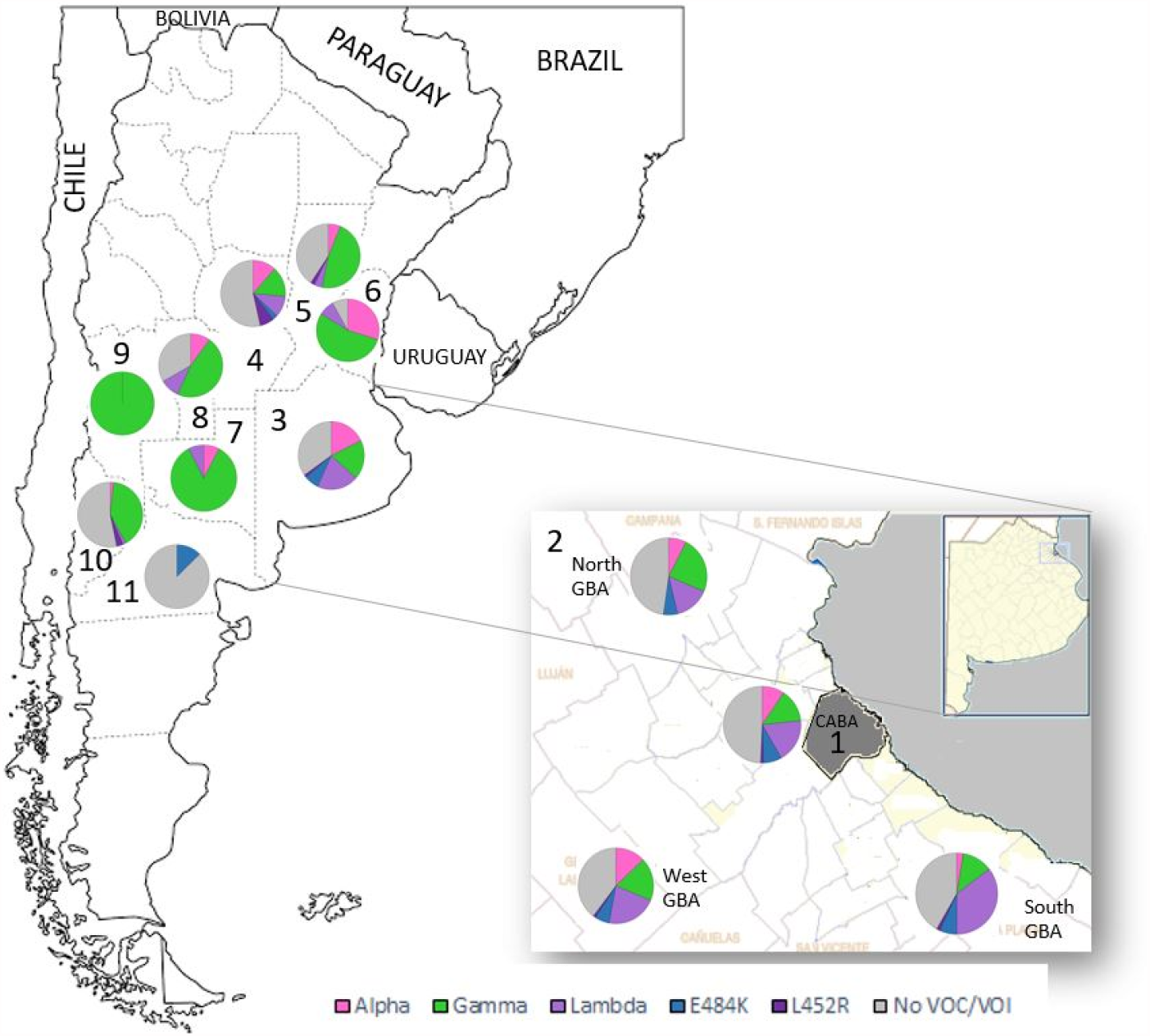
Location of cases analyzed in this work and pie charts representing the frequency of each variant detected in every region between EW44/2020 to EW20/2021. 1. Buenos Aires city, 2. Great Buenos Aires (North, West, South), 3. Province of Buenos Aires, 4. Province of Córdoba, 5. Province of Santa Fe, 6. Province of Entre Ríos, 7. Province of La Pampa, 8. Province of San Luis, 9. Province of Mendoza, 10. Province of Neuquén, 11. Province of Río Negro.

**Figure 2.**
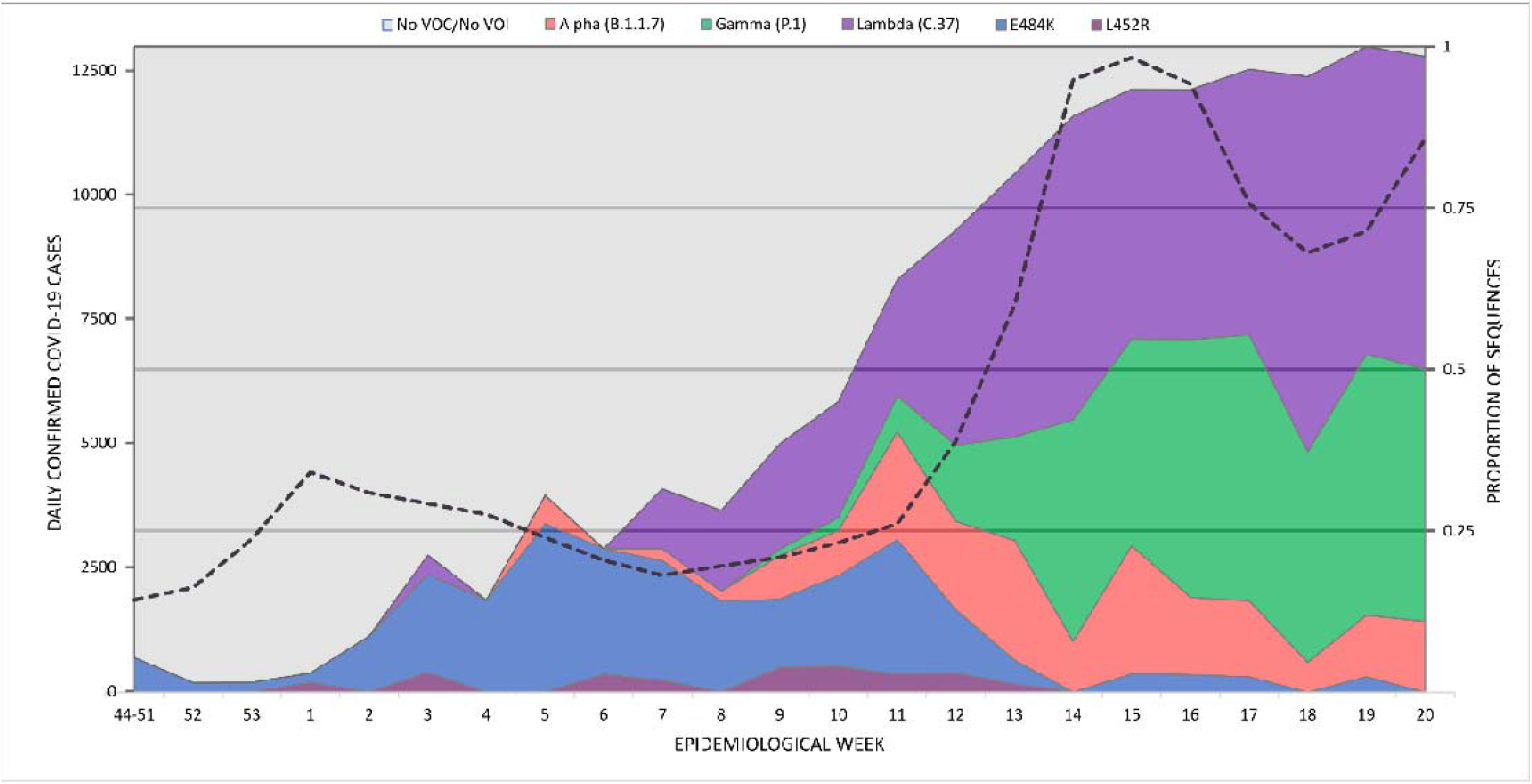
Number of reported cases by epidemiological week 2020-2021 since the beginning of the molecular surveillance of SARS-CoV-2 variants. Only cases that did not present a history of travel or close contact with travelers are included. Dotted line (mapped to the left y axis) indicates the average number of confirmed COVID-19 cases per week in the Metropolitan Area of Buenos Aires (MABA).

For this work, samples analyzed included a randomly selected 2.5-10% fraction of the total positive cases weekly detected in different health care centers. Regular sampling from four sentinel laboratories located in the metropolitan area of Buenos Aires (MABA) was performed along with sporadic sampling from other locations to sum up a total of 1,950 sequences. Surveillance strategy was based on Sanger sequencing of a 970bp region of Spike spanning amino acids 428 to 750 (Figure 3) (7). This region allows the identification of signature mutations associated with variants Alpha, Beta, Gamma, Lambda and Delta. Additionally, complete SARS-CoV-2 genome sequences were obtained from 456 samples using the Quick protocol (8) with Oxford Nanopore or Illumina platforms and combined with partial sequences to perform the analysis of 2,406 sequences (Table 1).

**Figure 3.**
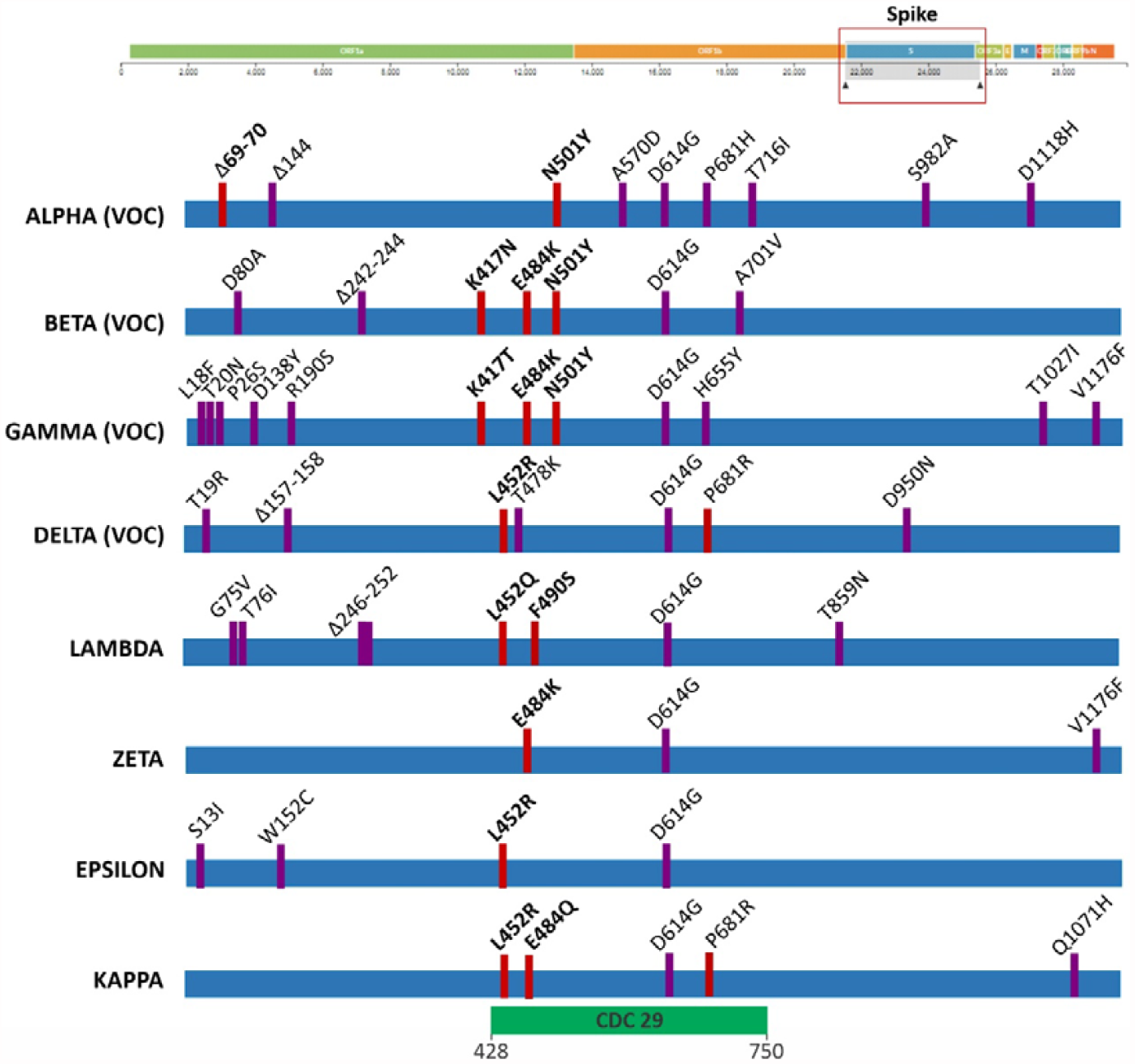
Representation of the amino acid changes in the S gene of SARS-CoV-2 variants of epidemiological interest. Changes indicated in red correspond to those with the greatest potential impact on viral biology or neutralization by antibodies. The location of CDC fragment 29 (codons 428 to 750) used for active surveillance of variants is indicated.

In this work we show genomic evidence of SARS-CoV-2 local transmission of variants Alpha, Gamma, Lambda, Epsilon and Zeta in Argentina, as well as the detection of mutations Spike_L452R and Spike_E484K in different geographic regions of the country.

**Alpha** was identified in 220 cases. This variant was detected in the city of Buenos Aires (CABA) and in the provinces of Buenos Aires, Córdoba, Entre Ríos, Santa Fe, San Luis, La Pampa and Neuquén. Its frequency in most of the MABA region (CABA plus Great Buenos Aires (GBA)) reached 19.8% (95% CI = 13.3-28.5) in EW 15/2021 (April 11^th^ to 17^th^) but decreased to 10.9% (95% CI = 5.1-21.2) in EW 20/2021 (May 16^th^ to 22^nd^) (Tables 1-2 and Figure 2). The phylogenetic analysis of whole genome sequences (including 127 from Argentina, 43 from this work) showed at least 35 independent introductions to the country, being the most related sequences from the USA, Central America, Europe, and the Middle East. Besides, five highly supported groups with at least three Argentinean sequences were observed (Figure 4). The largest group included 53 sequences from the CABA and nine provinces, from north to south of the country, suggesting a widespread local transmission and diversification.

**Figure 4.**
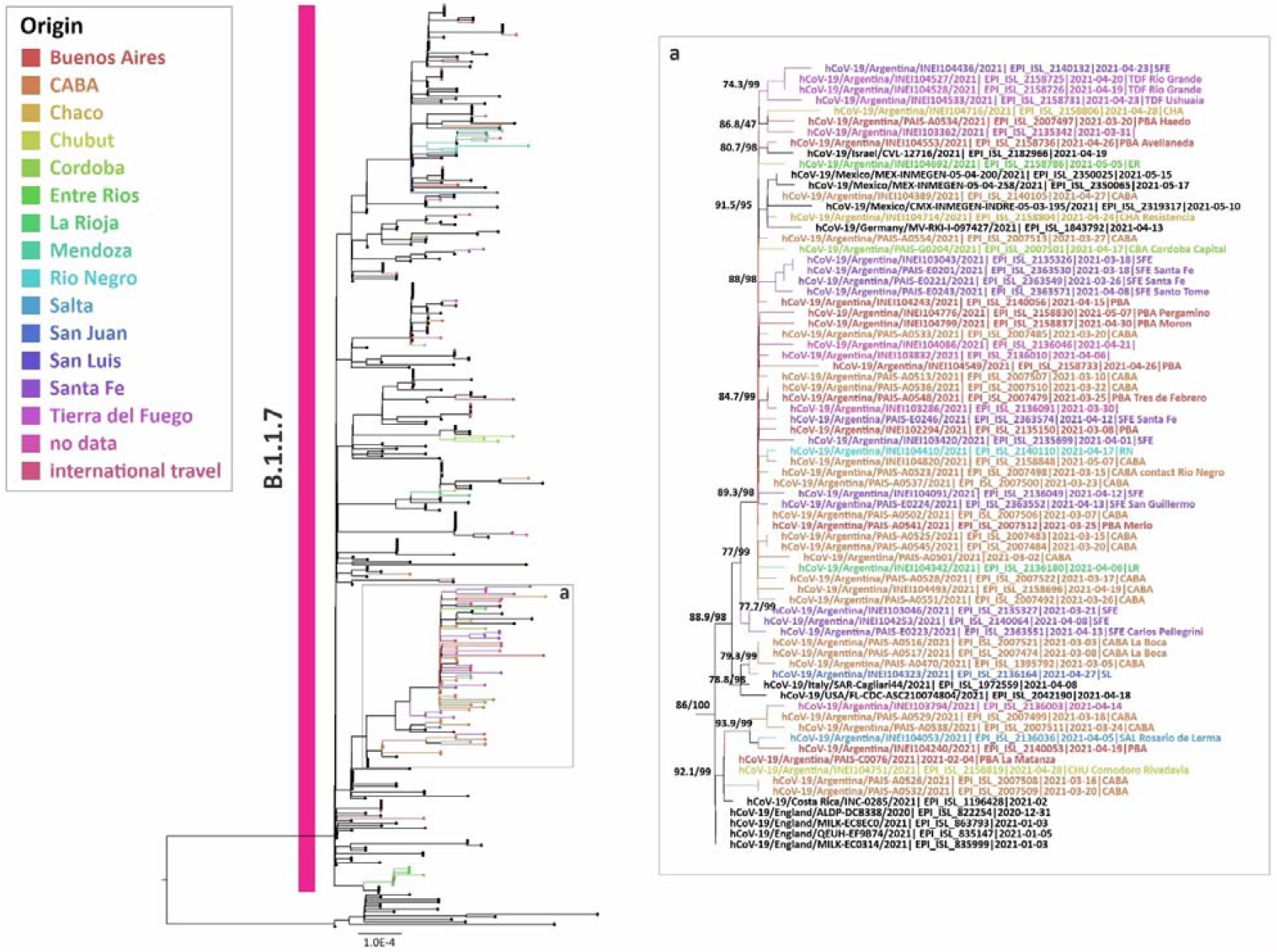
Phylogenetic tree of SARS-CoV-2 whole genome sequences of Alpha (lineage B.1.1.7). Dataset included sequences from Argentina, their best five BLAST hits sequences (against GISAID database on June 2^nd^ 313 2021), reference sequences of B.1.1.7 lineage, and B.1.1.1 sequences as outgroup. Only the largest group with Argentinean sequences is shown. Phylogenetic analysis was performed using IQ-TREE v. 2.1.2 COVID-edition (20). The S H-like approximate likelihood ratio test (1000 replicates) (21) and Ultrafast bootstrap Approximation (1000 replicates) (22) were used as methods to evaluate the reliability of the groups obtained. The SH-like / UFB values for the relevant groups are indicated for some groups. We gratefully acknowledge the authors from the originating laboratories responsible for obtaining the specimens and the submitting laboratories where genetic sequence data were generated and shared via the GISAID Initiative, on which part of this research is based (Table S1).

**Gamma** was identified in a total of 499 cases. This variant was detected in the CABA and in the provinces of Buenos Aires, Córdoba, Santa Fe, Entre Ríos, La Pampa, San Luis, Mendoza and Neuquén. Its frequency in the CABA and GBA remained stable at values over 30% since EW 16/2021, reaching 39.1% (95% CI = 28.0-51.3) in the EW 20/2021. The phylogenetic analysis of complete genome sequences (including 238 from Argentina, 50 from this work) showed at least 50 introductions to Argentina, with the most related sequences from Brazil and the USA. Besides, 18 supported groups with at least three Argentinean sequences were observed (Figure 5). The largest one included 72 sequences from the CABA and 14 provinces, from north to south of the country, also suggesting widespread local transmission and diversification (Figure 5.d). Eight sequences from Argentina were located in a separate clade within P.1 (previously named as P.1-like-II, (9)). This clade was formed by sequences from South America only (Brazil, Chile and Argentina) (Figure 5), showing a regional diversification process.

**Figure 5.**
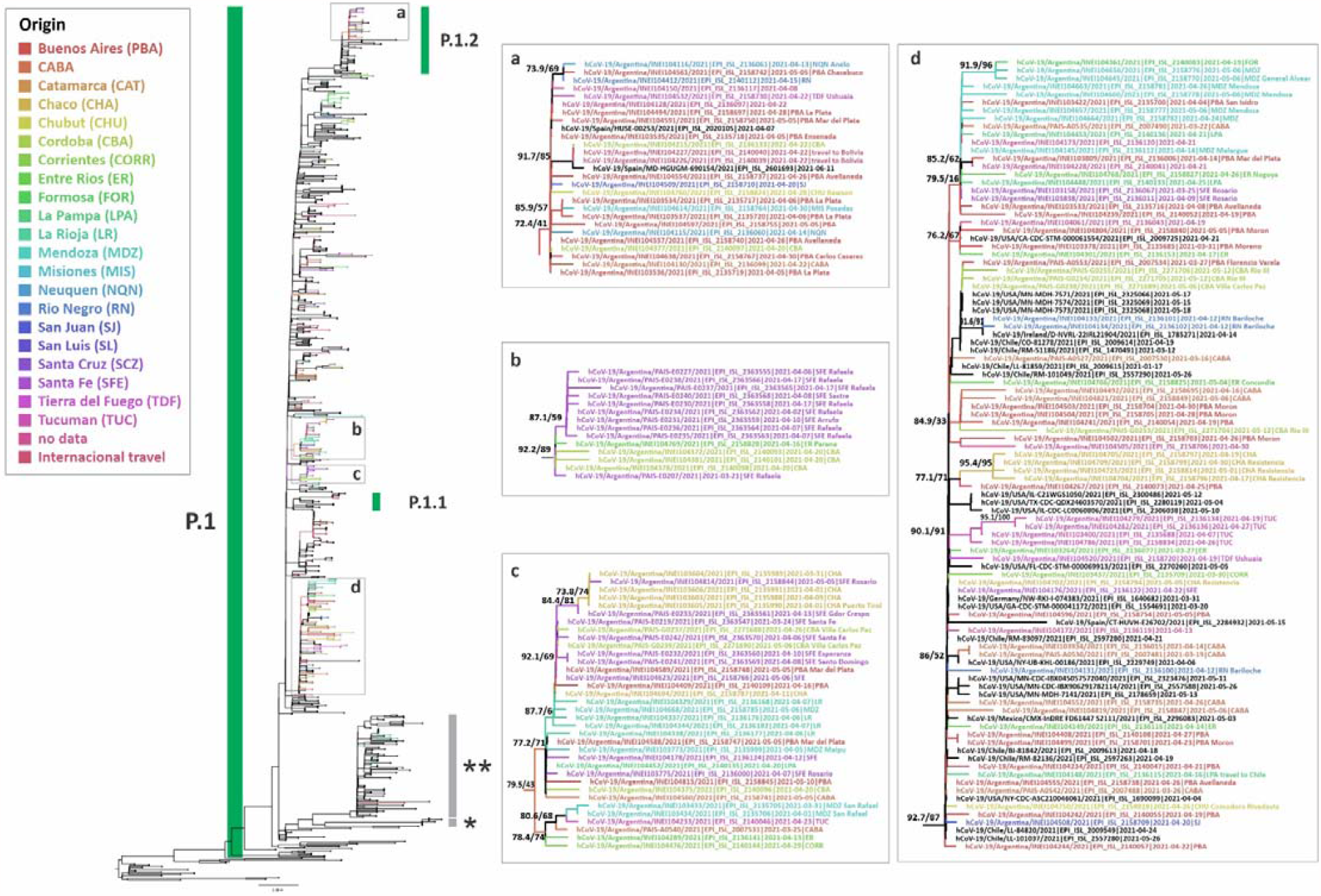
Phylogenetic tree of SARS-CoV-2 whole genome sequences of Gamma (lineage P.1). Dataset included sequences from Argentina, their best five BLAST hits sequences (against GISAID database on June 2^nd^ 321 2021), reference sequences of P.1 lineage, and B.1.1.28 sequences as outgroup. Only selected groups with Argentinean sequences are shown. Phylogenetic analysis was performed as previously described in Figure 5. *P.1-like-I and **P.1-like-II described by Gräf et al (9). We gratefully acknowledge the authors from the originating laboratories responsible for obtaining the specimens and the submitting laboratories where genetic sequence data were generated and shared via the GISAID Initiative, on which part of this research is based (Table S2).

**Lambda** variant showed a continuous increase since EW 7/2021 in the CABA and the GBA, reaching frequencies of 48.4% (95% CI = 36.6-60.4) in the EW 20/2021. Noteworthy, this variant appears to have replaced Alpha variant in the most populated region of the country (Table 2), given that since EW 15/2021, the proportion of cases associated with Alpha decreased while those associated with Lambda increased. These data have contributed to its recent declaration as a global VOI by the WHO (1). The transmission capacity, clinical behavior, and impact on vaccine effectiveness of this VOI will need further studies. However, preliminary studies have shown that the neutralization capacity of convalescent sera from the first wave viruses in Argentina and sera from individuals vaccinated with Sputnik V was not compromised (BBEI, 2021). On the other hand, increased infectivity and resistance to neutralizing antibodies produced by individuals immunized with CoronaVac (SinoVac) and mRNA-1273 (Moderna) vaccines was observed (10,11).

**Table 2:** Frequency of Alpha and Gamma, and mutations E484K and L452R by epidemiological week (EW) 2020-2021 in the Metropolitan Area of Buenos Aires (MABA).

| EW | Alpha |  | Gamma |  | Lambda |  | Mutation E484K<sup>3</sup> |  | Mutation L452R<sup>4</sup> |  | Non VOC/VOI |  | Total of sequences<sup>1</sup> |
| --- | --- | --- | --- | --- | --- | --- | --- | --- | --- | --- | --- | --- | --- |
|  | Frequency (%) | 95%CI<sup>2</sup> | Frequency (%) | 95%CI<sup>2</sup> | Frequency (%) | 95%CI<sup>2</sup> | Frequency (%) | 95%CI<sup>2</sup> | Frequency (%) | 95%CI<sup>2</sup> | Frequency (%) | 95%CI<sup>2</sup> |  |
| 44-51 | 0,0 | - | 0,0 | - | 0,0 | - | 5,4 | 2.0-12.3 | 0,0 | - | 94,6 | 87.7-98.0 | 93 |
| 52 | 0,0 | - | 0,0 | - | 0,0 | - | 1,5 | <0.01-8.6 | 0,0 | - | 98,5 | 91.4-100 | 68 |
| 53 | 0,0 | - | 0,0 | - | 0,0 | - | 1,5 | <0.01-8.9 | 0,0 | - | 98,5 | 91.1-100 | 66 |
| 1 | 0,0 | - | 0,0 | - | 0,0 | - | 1,5 | <0.01-8.8 | 1,5 | <0.01-8.8 | 97,0 | 89.1-99.8 | 67 |
| 2 | 0,0 | - | 0,0 | - | 0,0 | - | 8,7 | 2.9-20.9 | 0,0 | - | 91,3 | 79.1-97.1 | 46 |
| 3 | 0,0 | - | 0,0 | - | 3,0 | <0.01-16.7 | 15,2 | 6.2-31.4 | 3,0 | <0.01-16.7 | 78,8 | 62.0-89.6 | 33 |
| 4 | 0,0 | - | 0,0 | - | 0,0 | - | 14,3 | 5.8-29.9 | 0,0 | - | 85,7 | 70.1-94.2 | 35 |
| 5 | 4,4 | <0.01-22.7 | 0,0 | - | 0,0 | - | 26,1 | 12.3-46.8 | 0,0 | - | 69,6 | 48.9-84.6 | 23 |
| 6 | 0,0 | - | 0,0 | - | 0,0 | - | 19,4 | 9.5-35.3 | 2,8 | <0.01-15.4 | 77,8 | 61.7-88.5 | 36 |
| 7 | 1,9 | <0.01-10.7 | 0,0 | - | 9,3 | 3.6-20.3 | 18,5 | 10.2-31.0 | 1,9 | <0.01-10.7 | 68,5 | 55.2-79.4 | 54 |
| 8 | 1,6 | <0.01-9.1 | 0,0 | - | 12,5 | 6.2-23.0 | 14,1 | 7.4-24.8 | 0,0 | - | 71,9 | 59.8-81.5 | 64 |
| 9 | 6,7 | 3.1-13.5 | 1,0 | <0.01-5.8 | 16,4 | 10.4-24.7 | 10,6 | 5.9-18.1 | 3,9 | 1.2-9.8 | 61,5 | 51.9-70.3 | 104 |
| 10 | 7,0 | 3.2-14.0 | 2,0 | 0.1-7.4 | 18,0 | 11.6-26.8 | 14,0 | 8.4-22.3 | 4,0 | 1.2-10.2 | 55,0 | 45.2-64.4 | 100 |
| 11 | 16,7 | 9.6-27.1 | 5,6 | 1.8-13.8 | 18,1 | 10.7-28.6 | 20,8 | 12.9-31.7 | 2,8 | 0.2-10.2 | 36,1 | 26.0-47.7 | 72 |
| 12 | 13,7 | 8.2-21.9 | 11,8 | 6.7-19.6 | 33,3 | 24.9-43.0 | 9,8 | 5.2-17.3 | 2,9 | 0.6-8.7 | 28,4 | 20.6-37.9 | 102 |
| 13 | 18,5 | 11.4-28.5 | 16,1 | 9.5-25.7 | 40,7 | 30.7-51.6 | 3,7 | 0.8-10.8 | 1,2 | <0.01-7.3 | 19,8 | 12.4-29.8 | 81 |
| 14 | 7,8 | 3.8-14.9 | 34,3 | 25.8-44.0 | 47,1 | 37.7-56.7 | 0,0 | - | 0,0 | - | 10,8 | 6.0-18.4 | 102 |
| 15 | 19,8 | 13.3-28.5 | 32,1 | 23.9-41.5 | 38,7 | 30.0-48.2 | 2,8 | 0.6-8.4 | 0,0 | 37.0-65.0 | 6,6 | 3.0-13.2 | 106 |
| 16 | 12,0 | 6.2-21.5 | 40,0 | 29.7-51.3 | 38,7 | 28.4-50.0 | 2,7 | 0.2-9.8 | 0,0 | - | 6,7 | 2.5-15.0 | 75 |
| 17 | 11,8 | 6.3-20.5 | 41,2 | 31.3-51.8 | 41,2 | 31.3-51.8 | 2,4 | 0.1-8.7 | 0,0 | - | 3,5 | 0.8-10.3 | 85 |
| 18 | 4,7 | 0.4-16.3 | 32,6 | 20.4-47.6 | 58,1 | 43.3-71.6 | 0,0 | - | 0,0 | - | 4,7 | 0.4-16.3 | 43 |
| 19 | 9,5 | 3.2-22.6 | 40,5 | 27.0-55.5 | 47,6 | 33.4-62.3 | 2,4 | <0.01-13.4 | 0,0 | - | 0,0 | - | 42 |
| 20 | 10,9 | 5.1-21.2 | 39,1 | 28.0-51.3 | 48,4 | 36.6-60.4 | 0,0 | - | 0,0 | - | 1,6 | <0.01-9.1 | 64 |

<sup>1</sup> Only the cases from the MABA region that did not present a history of travel or close contact with travelers are included; in cases with a known epidemiological link between samples, only one was included in this table.
<sup>2</sup> The adjusted confidence interval of the frequency was estimated by the modified Wald method (19).
<sup>3</sup> Includes detections of the E484K mutation that do not belong to sequences with the characteristic combination of mutations of Gamma or Beta variants.
<sup>4</sup> Includes detections of the L452R mutation that do not belong to sequences with the characteristic combination of mutations of Delta or Kappa variants.

In addition, a probable case of coinfection of Alpha and Gamma was identified in a sample from the city of La Plata, and also two other cases of possible coinfection between Alpha and Lambda in the CABA. These cases will be further analyzed.

Importantly, the proportion of cases associated with Alpha and Gamma among individuals with no travel history or contact with travelers in CABA and GBA increased from less than 3% in the EW 7/2021-EW 9/2021 to 50.0% (for joint frequencies) in the EW 20/2021, and adding Lambda, they surpassed 98% of the total samples at that EW (Table 2).

However, the distribution of variants of epidemiological interest between EW 9/2021 and EW 20/2021 was heterogeneous within the MABA (Figure 6). While North GBA presented a predominance of Gamma (16/39 cases, 41.0%) and 9/39 cases of Lambda (23.1%), west GBA presented a more even distribution of variants, with Lambda in 58/176 cases (33.0%), Gamma in 49/176 cases (27.8%), and Alpha in 34/176 cases (19.3%). Similarly, the CABA presented Lambda in 150/508 cases (29.5%), Gamma in 110/508 cases (21.7%), and Alpha in 67/508 cases (13.2%). On the contrary, south GBA presented a predominance of Lambda (127/253 cases, 50.2%), followed by Gamma in 47/253 cases (18.6%) and Alpha in 10/253 cases (4.0%). Lastly, Great La Plata (GLP) showed a strong predominance of Gamma, counting 43/61 (70.5%).

**Figure 6.**
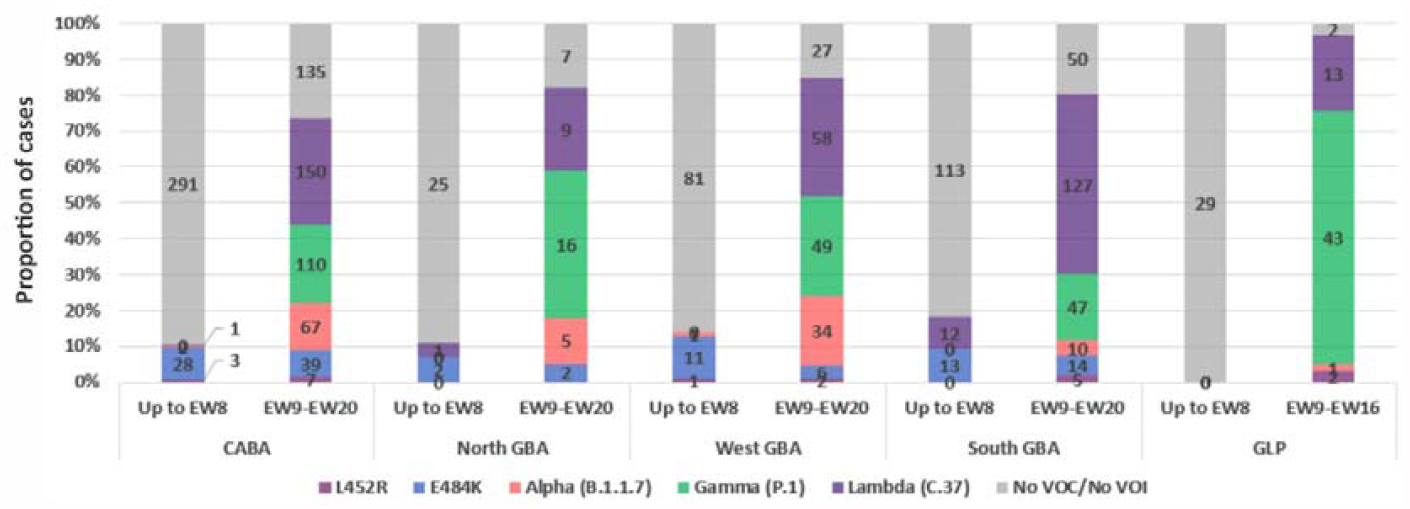
Cumulative number of SARS-CoV-2 variants and sequences with or without mutations of interest in the Metropolitan area of Buenos Aires (CABA, GBA and Great La Plata (GLP)). The cases were analyzed in two periods: until EW 8/2021 and from EW 9/2021 to EW 20/2021, according to the moment of change in the trend of the frequencies of the variants or mutations in each region. Only cases that did not present a history of travel or close contact with travelers are included; in cases sharing an epidemiological link, only one was considered as representative. The bars show the number of samples corresponding to variants, mutations or non-VOC/non-mutations of interest.

The analysis of the sporadic sampling from several places of the country also revealed a heterogeneous distribution of variants, showing a high proportion of cases associated with different variants and mutations of interest, especially with Gamma in most of the provinces studied (Table 1).

Therefore, the major lineages that circulated on the first wave at the beginning of the studied period were almost completely replaced by worldwide and regional emergent variants in a term of few weeks (Figure 2 and Figure 6), as was previously observed in other countries.

It is worth noting that data about the dynamics of the co-circulation of these highly transmissible variants are limited, given that in most countries only one of them became rapidly dominant and the introduction of a second VOC occurred after the first was already established. However, according to our data from several regions of Argentina, VOCs and VOIs presented similar frequencies in the population at the time of writing this report, which could allow proper comparative analyses of their dynamics, severity, and impact on vaccine effectiveness.

Mutations at aa positions 484 and 452 of Spike protein have been associated with possible immune escape and modified affinity to the human receptor and may occur in various newly emerging lineages worldwide (11–14). Both positions are located within the RBM (receptor binding motif). On one hand, the E484K mutation, constitutively present in Beta, Gamma, Zeta, Eta and Iota variants, was associated with resistance to neutralization by monoclonal antibodies, convalescent and vaccinated sera (4,12,15–17). On the other hand, mutations at position 452 of the Spike protein, were associated with decreased neutralization by monoclonal antibodies, convalescent and some vaccine sera (12,13). Another important mutation in Spike is P681R, associated with an enhanced viral fusion (18), which was not detected in this work.

The E484K mutation - not associated with Beta o Gamma signatures-was found in 138 cases. Most of the samples come from the CABA and the GBA regions from individuals without travel history, suggesting local circulation, at least, since the last week of December 2020, when it was first detected by our surveillance program. Nevertheless, a reduction in its frequency was observed since EW 12/2021 in the CABA and the GBA, being almost completely replaced by VOCs and VOIs of more recent emergence, such as Lambda (Table 2). So far, 53 of 138 cases analyzed by full-length sequencing have been identified as belonging to the Zeta variant through the analysis of the complete genome.

For the L452R mutation, a similar replacement has been observed, being occasionally detected in the CABA and the GBA between the EW 1/2021 and the EW 13/2021, but not thereafter. The complete genome analysis of 21 cases allowed to identify the Epsilon variant in 18 cases: one from lineage B.1.429 and 17 from lineage B.1.427. None of the L452R cases detected so far were associated with Delta or Kappa variants.

In conclusion, the surveillance strategy implemented over 30 epidemiological weeks in Argentina, based on Spike and complete genome sequencing, allowed to describe the introduction, establishment, and evolution of SARS-CoV-2 variants of interest and concern in the second wave of the COVID-19 pandemic in South America. The main variants found were Gamma, Lambda and Alpha, in that order, with lower detection of Zeta and Epsilon. This implementation allowed a rapid response in a limited resource access scenario and contributed to the implementation of public health measures to control disease spread.

## Data Availability

The sequences obtained in this study can be found in the online repository GISAID.

