## Supplementary material for "Surveillance of SARS-CoV-2 variants in Argentina: detection of Alpha, Gamma, Lambda, Epsilon and Zeta in locally transmitted and imported cases": Table S1

We gratefully acknowledge the following Authors from the Originating laboratories responsible for obtaining the specimens, as well as the Submitting laboratories where the genome data were generated and shared via GISAID, on which this research is based.

All Submitters of data may be contacted directly via www.gisaid.org

Authors are sorted alphabetically.

| Accession ID | Originating Laboratory | Submitting Laboratory | Authors |
|---|---|---|---|
| EPI_ISL_1013610 | Department for Molecular Diagnostics, Centre for Medical Microbiology, Institute of Public Health, Montenegro | Charité Universitätsmedizin Berlin, Institut für Virologie | Victor M Corman, Barbara Mühlemann, Jörn Beheim-Schwarzbach, Julia Tesch, Tobias Bleicker, Danijela Vujoševi, Marija Govedarica, Talitha Veith, Julia Schneider, Terry Jones, Christian Drosten |
| EPI_ISL_1051869 | Oxford Viromics, NDM, University of Oxford; Oxford University Hospitals; Basingstoke and North Hampshire Hospital | COVID-19 Genomics UK (COG-UK) Consortium | Tanya Golubchik, David Bonsall, George Macintyre, Amy Trebes, Mariateresa de Cesare, Catrin Moore, Alex Mobbs, Anita Justice, Robert Shaw, Monique Andersson, Timothy Peto, Emma Wise, Nathan Moore, Jessica Lynch, Nick Cortes, Matilde Mori, Stephen Kidd, David Buck, John Todd, Christophe Fraser |
| EPI_ISL_1088294 | Quest Diagnostics Incorporated | Respiratory Viruses Branch, Division of Viral Diseases, Centers for Disease Control and Prevention | Peter W. Cook, Dakota Howard, Dhwani Batra, Ben L. Rambo-Martin, S. H. Rosenthal, A. Gerasimova, R. M. Kagan, B. Anderson, M. Hua, Y. Liu, L.E. Bernstein, K.E. Livingston, A. Perez, I. A. Shlyakhter, R. V. Rolando, R. Owen, P. Tanpaiboon, F. Lacbawan, Clinton R. Paden, Suxiang Tong, Duncan MacCannell |
| EPI_ISL_1092009 | Kansas Health and Environmental Lab | Kansas Health and Environmental Lab | Mike Grose, Paige Drury, Carissa Robertson, Ben Olsen, and Phil Adam |
| EPI_ISL_1093172 | Laboratorio de Referencia Nacional de Virus Respiratorio. Instituto Nacional de Salud Perú | Laboratorio de Referencia Nacional de Biotecnología y Biología Molecular. Instituto Nacional de Salud Perú | Carlos Padilla Rojas, Karolyn Vega Chozo, Luis Barcena, Priscila Lope Pari, Omar Caceres Rey, Marco Galarza Perez, Maribel Huaringa Nuñez, Johanna Balbuena Torrez, Henri Bailon Calderon, Nancy Rojas Serrano |
| EPI_ISL_1150267 | Sonic - Labor Dr. von Froreich GmbH | Robert Koch Institute | unknown |
| EPI_ISL_1172872 | Pandemic Response Lab - NYC | Pandemic Response Lab, R&D | Henry Lee, Michael Hammerling, Melissa Hopkins, Cybill del Castillo, Shinyoung Clair Kang, William Ward, Pradeep Bugga, Haiping Hao, Jon Laurent |
| EPI_ISL_1179460 | Originating lab: Wales Specialist Virology Centre Sequencing lab: Pathogen Genomics Unit | Public Health Wales Microbiology Cardiff Wales Specialist Virology Centre | Catherine Moore, Johnathan Evans, Laura Gifford, Malorie Perry, Simon Cottrell, Angela Marchbank, Alec Birchley, Alexander Adams, Amy Gaskin, Bree Gatica-Wilcox, Jason Coombes, Joel Southgate, Lauren Gilbert, Lee Graham, Nicole Pacchiarini, Sara Kumziene-Summerhayes, Sarah Taylor, Sophie Jones, Sara Rey, Matthew Bull, Joanne Watkins, Sally Corden, Tom Connor |
| EPI_ISL_1196428 | CLINICA BIBLICA | Inciensa, Instituto Costarricense de Investigación y Enseñanza en Nutrición y Salud | Francisco Duarte, Hebleen Porras, Claudio Soto-Garita, Estela Cordero, Adriana Godínez, Melany Calderón & Karla Gutiérrez-González |
| EPI_ISL_1240208 | Jessa | Jessa | Cruys et al. on behalf of the Jessa_cmdLab |
| EPI_ISL_1249076 | Oxford Viromics, NDM, University of Oxford; Oxford University Hospitals; Basingstoke and North Hampshire Hospital | COVID-19 Genomics UK (COG-UK) Consortium | Tanya Golubchik, David Bonsall, George Macintyre, Amy Trebes, Mariateresa de Cesare, Catrin Moore, Alex Mobbs, Anita Justice, Robert Shaw, Monique Andersson, Timothy Peto, Emma Wise, Nathan Moore, Jessica Lynch, Nick Cortes, Matilde Mori, Stephen Kidd, David Buck, John Todd, Christophe Fraser |
| EPI_ISL_1284639 | Sonic - Labor Dr. von Froreich GmbH | Robert Koch Institute | unknown |
| EPI_ISL_1297282, EPI_ISL_1297306 | BIOMNIS PARIS | CNR Virus des Infections Respiratoires - France SUD | Antonin Bal, Gregory Destras, Gwendolyne Burfin, Hadrien Regue, Quentin Semanas, Martine Valette, Bruno Lina, Laurence Josset |
| EPI_ISL_1313062 | BIOMNIS LYON | CNR Virus des Infections Respiratoires - France SUD | Antonin Bal, Gregory Destras, Gwendolyne Burfin, Hadrien Regue, Quentin Semanas, Martine Valette, Bruno Lina, Laurence Josset |
| EPI_ISL_1322212 | IrsiCaixa | IrsiCaixa | Marc Noguera-Julian, Mariona Parera, Maria Casadellà, Pilar Armengol, Francesc Catala-Moll, Roger Paredes, Bonaventura Clotet Gloria Trujillo, Rafel Perez Vidal, Jaume Trape Pujol, Carolina Gonzalez Fernandez, Roger Paredes, Eulalia Grau, Bonaventura Clotet |
| EPI_ISL_1337155 | Helix/Illumina | Centers for Disease Control and Prevention Division of Viral Diseases, Pathogen Discovery | Peter W. Cook, Dakota Howard, Dhwani Batra, Ben L. Rambo-Martin, Eileen de Feo, Jan Antico, Christine Tran, Matthew Tolentino, Shannon Wickline, Kim Gietzen, Brad Sickler, Jingtao Liu, Eric Allen, Phil Febbo, Summer Galloway, Nicole L. Washington, Simon White, Geraint Levan, Kelly Schiabor Barrett, Elizabeth Cirulli, Alexandre Bolze, Ary Ascencio, Charlotte Rivera-Garcia, Ryan Cho, Jason Nguyen, Sherry Wang, Jimmy Ramirez, Tyler Cassens, Efren Sandoval, Magnus Isaksson, William Lee, David Becker, Marc Laurent, James Lu, Clinton R. Paden, Suxiang Tong, Duncan MacCannell |
| EPI_ISL_1355284 | Universitätsklinikum Leipzig - Institut für Medizinische Mikrobiologie und Virologie virologisches Labor | Robert Koch Institute | unknown |
| EPI_ISL_1360284 | Hospital Universitari Bellvitge | Microbiology Department | Sara Marti, Aida Gonzalez-Diaz, Laura Calatayud, Jordi Niubó, Miguel Fernandez-Huerta, Carmen Ardanuy, Jordi Camara, M Angeles Domínguez |
| EPI_ISL_1394456 | Hospital Universitari Vall d'Hebron - Vall d'Hebron Institut de Recerca | Hospital Universitari Vall d'Hebron - Vall d'Hebron Institut de Recerca | Cristina Andrés, Maria Piñana, Josep F Abril, Damir Garcia-Cehic, Ariadna Rando, Juliana Esperalba, Maria Gema Codina, Carla Castillo, Maria Carmen Martin, Tomàs Pumarola, Josep Quer, Andrés Antón |
| EPI_ISL_1395792 | Laboratorio de Virología del Hospital de Niños Dr. Ricardo Gutierrez | Área de Secuenciación del Laboratorio de Virología del Hospital de Niños Dr. Ricardo Gutierrez on behalf of 'Proyecto Argentino Interinstitucional de genomica de SARS-CoV-2' (PAIS Consortium) | Alexay, S; Thomas, G; Medina, C; Labarta, N; Streitenberger, C; Villegas, E; Barreda Frank, M; Grandis, E; Acevedo, ME; Alvarez Lopez, C; Jacques, O; Mistchenko, A; Nabaes Jodar, M; Goya, S; Lusso, S; Acuña, D; Natale, MI; Valinotto, LE; Viegas, M. |
| EPI_ISL_1402899 | Johns Hopkins Hospital Department of Pathology | Johns Hopkins Hospital Department of Pathology | C. Paul Morris, Chun Huai Luo, Adannaya Amadi, Matthew Schwartz, Heba H. Mostafa |
| EPI_ISL_1406037, EPI_ISL_1406062 | Microbiology Department, Laboratori Clínic Metropolitana Nord. Hospital Universitari Germans Trias i Pujol. | Can Ruti SARS-CoV-2 Sequencing Hub (HUGTiP/IrsiCaixa/IGTP) | Marc Noguera-Julian, Pilar Armengol, Ignacio Blanco, Antoni E Bordoy, Francesc Catala-Moll, Pere-Joan Cardona, Maria Casadellà, Cristina Casañ, Gemma Clara, Bonaventura Clotet, Cristina Esteban, Montserrat Giménez, Mercedes Guerrero, Anna Not, Roger Paredes, Mariona Parera, Verónica Saludes, Alba Sánchez, and Elisa Martró on behalf of the Can Ruti SARS-CoV-2 Sequencing Hub. |
| EPI_ISL_1465944 | Lighthouse Lab in Cambridge | Wellcome Sanger Institute for the COVID-19 Genomics UK (COG-UK) Consortium | Rob Howes, The Lighthouse Lab in Cambridge and Alex Alderton, Roberto Amato, Jeffrey Barrett, Sonia Goncalves, Ewan Harrison, David K. Jackson, Ian Johnston, Dominic Kwiatkowski, Cordelia Langford, John Sillitoe on behalf of the Wellcome Sanger Institute COVID-19 Surveillance Team |
| EPI_ISL_1468575 | Johns Hopkins Hospital Department of Pathology | Johns Hopkins Hospital Department of Pathology | C. Paul Morris, Chun Huai Luo, Adannaya Amadi, Matthew Schwartz, Heba H. Mostafa |
| EPI_ISL_1471732 | Pandemic Response Lab - NYC | Pandemic Response Lab, R&D | Henry Lee, Michael Hammerling, Melissa Hopkins, Cybill del Castillo, Shinyoung Clair Kang, William Ward, Pradeep Bugga, Sol Rey, Dylan Law, Haiping Hao, Jon Laurent |
| EPI_ISL_1503243 | Wichita State University - Molecular Diagnostics Lab | Kansas Health and Environmental Lab | Mike Grose, Jonathan Barnell, Ben Olsen, and Phil Adam |
| EPI_ISL_1524827 | Hospital General Universitario Gregorio Marañón | Hospital General Universitario Gregorio Marañón | Sergio Buenestado Serrano, Pedro Sola Campoy, Laura Pérez-Lago, Cristina Rodriguez-Grande, Pilar Catalán, Patricia Muñoz, Darío García de Viedma |
| EPI_ISL_1528573 | WVU Rapid Development Lab | WVU and Marshall University Combined Genomics Core Facilities | "James Denvir, Peter Stoilov, Peter Perrotta, Wesley Kimble, Ryan Percifield" |
| EPI_ISL_1543279 | Pandemic Response Lab - NYC | Pandemic Response Lab, R&D | Henry Lee, Michael Hammerling, Melissa Hopkins, Cybill del Castillo, Shinyoung Clair Kang, William Ward, Pradeep Bugga, Sol Rey, Dylan Law, Katharine Nelson, Haiping Hao, Jon Laurent |
| EPI_ISL_1550427, EPI_ISL_1560274, EPI_ISL_1560525 | Aegis Sciences Corporation | Centers for Disease Control and Prevention Division of Viral Diseases, Pathogen Discovery | Dakota Howard, Dhwani Batra, Peter W. Cook, Kara Moser, Adrian Paskey, Jason Caravas, Benjamin Rambo-Martin, Shatavia Morrison, Christopher Gulvick, Scott Sammons, Yvette Unoarumhi, Darlene Wagner, Matthew Schmerer, Cyndi Clark, Patrick Campbell, Rob Case, Vikramsinha Ghorpade, Holly Houdeshell, Ola Kvalvaag, Dillon Nall, Ethan Sanders, Alec Vest, Shaun Westlund, Matthew Hardison, Clinton R. Paden, Duncan MacCannell |
| EPI_ISL_1575599 | Helix/Illumina | Centers for Disease Control and Prevention Division of Viral Diseases, Pathogen Discovery | Dakota Howard, Dhwani Batra, Peter W. Cook, Kara Moser, Adrian Paskey, Jason Caravas, Benjamin Rambo-Martin, Shatavia Morrison, Christopher Gulvick, Scott Sammons, Yvette Unoarumhi, Darlene Wagner, Matthew Schmerer, Eileen de Feo, Jan Antico, Christine Tran, Matthew Tolentino, Shannon |

| | | | |
|---|---|---|---|
| | | | Wickline, Kim Gietzen, Brad Sickler, Jingtao Liu, Eric Allen, Phil Febbo, Nicole L. Washington, Simon White, Geraint Levan, Kelly Schiabor Barrett, Elizabeth Cirulli, Alexandre Bolze, Ary Ascencio, Charlotte Rivera-Garcia, Ryan Cho, Jason Nguyen, Sherry Wang, Jimmy Ramirez, Tyler Cassens, Efren Sandoval, Magnus Isaksson, William Lee, David Becker, Marc Laurent, James Lu, Clinton R. Paden, Duncan MacCannell |
| EPI_ISL_1623327 | NOVABIO DORDOGNE | CNR Virus des Infections Respiratoires - France SUD | Antonin Bal, Gregory Destras, Gwendolyne Burfin, Hadrien Regue, Quentin Semanas, Martine Valette, Bruno Lina, Laurence Josset |
| EPI_ISL_1667474 | Department of Laboratory Medicine, National Taiwan University Hospital | Microbial Genomics Core Lab, National Taiwan University Centers of Genomic and Precision Medicine | Shiou-Hwei Yeh, You-Yu Lin, Ya-Yun Lai, Chiao-Ling Li, Shan-Chwen Chang, Pei-Jer Chen, Sui-Yuan Chang |
| EPI_ISL_1669359 | Hospital Universitari Vall d'Hebron - Vall d'Hebron Institut de Recerca | Hospital Universitari Vall d'Hebron - Vall d'Hebron Institut de Recerca | Cristina Andrés, Maria Piñana, Damir Garcia-Cehic, Ariadna Rando, Juliana Esperalba, Maria Gema Codina, Carla Castillo, Maria Carmen Martin, Tomàs Pumarola, Josep Quer, Andrés Antón |
| EPI_ISL_1671609 | NORTHWELL HEALTH LABORATORIES | Wadsworth Center, New York State Department of Health | Kirsten St. George, Daryl M. Lamson, Alexis Russell, Matthew Shudt, Melissa A Leisner, Jonathan Plitnick, Catharine Prussing, Navjot Singh, John Kelly, Erasmus Schneider, Erica Lasek-Nesselquist |
| EPI_ISL_1682599, EPI_ISL_1683246 | Laboratory Corporation of America | Centers for Disease Control and Prevention Division of Viral Diseases, Pathogen Discovery | Dakota Howard, Dhwani Batra, Peter W. Cook, Kara Moser, Adrian Paskey, Jason Caravas, Benjamin Rambo-Martin, Shatavia Morrison, Christopher Gulvick, Scott Sammons, Yvette Unoarumhi, Darlene Wagner, Matthew Schmerer, Minoo Agarwal, Eyad Almasri, Debbie Boles, Ayla Burns, Nuthawin Charoensri, Oren Cohen, Susan Countryman, Mary Ann Cristobal, Bobbi Croy, Suzanne Dale, Hrushikesh Deshmukh, Amanda Douglas, Vincent Drouillon, Marcia Eisenberg, Howard Engler, Rama Ghatti, Prashant Gupta, Susan Hicks, Jake Humphrey, Lax Iyer, Manoj Jain, Mohan Kolli, Brian Krueger, Tim Kuphal, Stanley Letovsky, Michael Levandoski, Craig Lukasik, Jonathan Meltzer, Brian Norvell, Mindy Nye, Scott Parker, Christos Petropoulos, John Pruitt, Steven Ragan, Scott Ryan, Mike Sapeta, Jana Schroth, Suresh Babu Selvaraju, Goran Stevovic, Amanda Suchanek, Andrea Throop, Lyndon Tilson, Thomas Urban, Joe Voshell, Kimberly Wagner, Jonathan Williams, Mary Williamson, Qian Zeng, Tricia Zwiefelhofer, Clinton R. Paden, Duncan MacCannell |
| EPI_ISL_1688958, EPI_ISL_1689915, EPI_ISL_1690014, EPI_ISL_1690016 | Aegis Sciences Corporation | Centers for Disease Control and Prevention Division of Viral Diseases, Pathogen Discovery | Dakota Howard, Dhwani Batra, Peter W. Cook, Kara Moser, Adrian Paskey, Jason Caravas, Benjamin Rambo-Martin, Shatavia Morrison, Christopher Gulvick, Scott Sammons, Yvette Unoarumhi, Darlene Wagner, Matthew Schmerer, Cyndi Clark, Patrick Campbell, Rob Case, Vikramsinha Ghorpade, Holly Houdeshell, Ola Kvalvaag, Dillon Nall, Ethan Sanders, Alec Vest, Shaun Westlund, Matthew Hardison, Clinton R. Paden, Duncan MacCannell |
| EPI_ISL_1694150, EPI_ISL_1694225 | Infinity Biologix | Centers for Disease Control and Prevention Division of Viral Diseases, Pathogen Discovery | Dakota Howard, Dhwani Batra, Peter W. Cook, Kara Moser, Adrian Paskey, Jason Caravas, Benjamin Rambo-Martin, Shatavia Morrison, Christopher Gulvick, Scott Sammons, Yvette Unoarumhi, Darlene Wagner, Matthew Schmerer, Christian Bixby, Yihe Wang, Jonathan Schultz, Chirayu Goswami, Russ Hager, Robin Grimwood, Clinton R. Paden, Duncan MacCannell |
| EPI_ISL_1728276 | SYNLAB MVZ Leverkusen | Robert Koch Institute | unknown |
| EPI_ISL_1737004 | Aegis Sciences Corporation | Centers for Disease Control and Prevention Division of Viral Diseases, Pathogen Discovery | Dakota Howard, Dhwani Batra, Peter W. Cook, Kara Moser, Adrian Paskey, Jason Caravas, Benjamin Rambo-Martin, Shatavia Morrison, Christopher Gulvick, Scott Sammons, Yvette Unoarumhi, Darlene Wagner, Matthew Schmerer, Cyndi Clark, Patrick Campbell, Rob Case, Vikramsinha Ghorpade, Holly Houdeshell, Ola Kvalvaag, Dillon Nall, Ethan Sanders, Alec Vest, Shaun Westlund, Matthew Hardison, Clinton R. Paden, Duncan MacCannell |
| EPI_ISL_1805222 | Maryland Genomics, Institute for Genome Sciences, University of Maryland School of Medicine | Maryland Genomics, Institute for Genome Sciences, University of Maryland School of Medicine | Tallon, Luke J; Sadzewicz, Lisa D; Humphrys, Mike; Ott, Sandra; Roussey, Holly; Mehta, Aditya; Vavikolanu, Kranthi; Fraser, Claire M; Ravel, Jacques |
| EPI_ISL_1807213 | GA Department of Public Health | GA Department of Public Health | Stacy Reeves, Jonathan Edwards, Cynthia Dixey, Tonia Parrott, Aliyah Fields, Taylor Smith |
| EPI_ISL_1836219, EPI_ISL_1838360 | Aegis Sciences Corporation | Centers for Disease Control and Prevention Division of Viral Diseases, Pathogen Discovery | Dakota Howard, Dhwani Batra, Peter W. Cook, Kara Moser, Adrian Paskey, Jason Caravas, Benjamin Rambo-Martin, Shatavia Morrison, Christopher Gulvick, Scott Sammons, Yvette Unoarumhi, Darlene Wagner, Matthew Schmerer, Cyndi Clark, Patrick Campbell, Rob Case, Vikramsinha Ghorpade, Holly Houdeshell, Ola Kvalvaag, Dillon Nall, Ethan Sanders, Alec Vest, Shaun Westlund, Matthew Hardison, Clinton R. Paden, Duncan MacCannell |
| EPI_ISL_1841485, EPI_ISL_1841520, EPI_ISL_1841707 | Department of Virology and Immunology, University of Helsinki and Helsinki University Hospital, Huslab Finland | Department of Virology, Faculty of Medicine, University of Helsinki, Helsinki, Finland | Teemu Smura, Ravi Kant, Phuoc Truong, Hussein Alburkat, Hannimari Kallio-Kokko, Jenni Virtanen, Maija Suvanto, Essi Korhonen, Sari Hannula, Harri Kangas, Hanna Liimatainen, Satu Kurkela, Hanna Jarva, Maija Lappalainen, Pekka Ellonen, Olli Vapalahti |
| EPI_ISL_1843792 | Centogene; Dr. Bauer Laboratoriums GmbH | Robert Koch Institute | unknown |
| EPI_ISL_1844122 | Bioscientia Labor Wermsdorf | Robert Koch Institute | unknown |
| EPI_ISL_1844532 | IMD - MVZ Labor Martinsried | Robert Koch Institute | unknown |
| EPI_ISL_1882007 | Department of Virus and Microbiological Special Diagnostics, Statens Serum Institut, Copenhagen, Denmark | Aalborg University | Danish Covid-19 Genome Consortium |
| EPI_ISL_1922367 | Servicio de Microbiología Clínica (Complejo Hospitalario de Navarra, Pamplona) | Centro de Secuenciación NASERTIC | Carmen Ezpeleta Baquedano, Ana Navascués, Ana Miqueleiz |
| EPI_ISL_1925276 | Aegis Sciences Corporation | Centers for Disease Control and Prevention Division of Viral Diseases, Pathogen Discovery | Dakota Howard, Dhwani Batra, Peter W. Cook, Kara Moser, Adrian Paskey, Jason Caravas, Benjamin Rambo-Martin, Shatavia Morrison, Christopher Gulvick, Scott Sammons, Yvette Unoarumhi, Darlene Wagner, Matthew Schmerer, Cyndi Clark, Patrick Campbell, Rob Case, Vikramsinha Ghorpade, Holly Houdeshell, Ola Kvalvaag, Dillon Nall, Ethan Sanders, Alec Vest, Shaun Westlund, Matthew Hardison, Clinton R. Paden, Duncan MacCannell |
| EPI_ISL_1964068, EPI_ISL_1964073 | Hospital Universitari Bellvitge | Microbiology Department | Sara Marti, Aida Gonzalez-Diaz, Laura Calatayud, Jordi Niubó, Miguel Fernandez-Huerta, Carmen Ardanuy, Jordi Camara, M Angeles Domínguez |
| EPI_ISL_1971160 | Weill Cornell Medicine | New York Genome Center | Michael Zody, Andre Corvelo, Dayna M. Oschwald, Samantha Fennessey, Tom Maniatis, Melissa Cushing, Olivier Elemento, Margaret Elizabeth Ross, Chris Mason, Priya Velu, Hanna Rennert, Arryn Craney, Lars F Westblade |
| EPI_ISL_1972559 | Laboratorio HUB -Azienda Ospedaliero Universitaria - AOU - Cagliari | Laboratorio SPOKE Biologia Molecolare -Azienda Ospedaliero Universitaria - AOU - Cagliari | Germano Orrù, Sara Fais, Valentina Medda, Alessandra Scano, Miriam Loddo, Riccardo Cappai, Ferdinando Coghe |
| EPI_ISL_1991693 | Helix/Illumina | Centers for Disease Control and Prevention Division of Viral Diseases, Pathogen Discovery | Dakota Howard, Dhwani Batra, Peter W. Cook, Kara Moser, Adrian Paskey, Jason Caravas, Benjamin Rambo-Martin, Shatavia Morrison, Christopher Gulvick, Scott Sammons, Yvette Unoarumhi, Darlene Wagner, Matthew Schmerer, Eileen de Feo, Jan Antico, Christine Tran, Matthew Tolentino, Shannon Wickline, Kim Gietzen, Brad Sickler, Jingtao Liu, Eric Allen, Phil Febbo, Nicole L. Washington, Simon White, Geraint Levan, Kelly Schiabor Barrett, Elizabeth Cirulli, Alexandre Bolze, Ary Ascencio, Charlotte Rivera-Garcia, Ryan Cho, Jason Nguyen, Sherry Wang, Jimmy Ramirez, Tyler Cassens, Efren Sandoval, Magnus Isaksson, William Lee, David Becker, Marc Laurent, James Lu, Clinton R. Paden, Duncan MacCannell |
| EPI_ISL_1995941, EPI_ISL_1996423, EPI_ISL_1996557, EPI_ISL_2000300 | Aegis Sciences Corporation | Centers for Disease Control and Prevention Division of Viral Diseases, Pathogen Discovery | Dakota Howard, Dhwani Batra, Peter W. Cook, Kara Moser, Adrian Paskey, Jason Caravas, Benjamin Rambo-Martin, Shatavia Morrison, Christopher Gulvick, Scott Sammons, Yvette Unoarumhi, Darlene Wagner, Matthew Schmerer, Cyndi Clark, Patrick Campbell, Rob Case, Vikramsinha Ghorpade, Holly Houdeshell, Ola Kvalvaag, Dillon Nall, Ethan Sanders, Alec Vest, Shaun Westlund, Matthew Hardison, Clinton R. Paden, Duncan MacCannell |
| EPI_ISL_2003803, EPI_ISL_2003928 | Hospital of the University of Pennsylvania Molecular Pathology Lab | Bushman Lab - University of Pennsylvania | John Everett, Kyle Rodino, Shantan Reddy, Pascha Hokama, Aoife M. Roche, Young Hwang, Abigail Glascock, Scott Sherrill-Mix, Samantha A. Whiteside, Jevon Graham-Wooten, Layla A. Khatib, Ayannah S. Fitzgerald, Arupa Ganguly, Mike Feldman, Brendan Kelly, Ronald G. Collman and Frederic Bushman |
| EPI_ISL_2007474 | Hospital General de Agudos Dr. Cosme Argerich | Área de Secuenciación del Laboratorio de Virología del Hospital de Niños Dr. Ricardo Gutierrez on behalf of 'Proyecto Argentino Interinstitucional de genomica de SARS-CoV-2' (PAIS Consortium) | Marcia Pozzati, Jésica Galeano, Florencia Rodríguez, Florencia Funez, Andrea Fernández, Karina Polanski; Alexay, S; Nabaes Jodar, M; Acuña, D; Goya, S; Lusso, S; Natale, MI; Valinotto, LE; Viegas, M. |
| EPI_ISL_2007479, EPI_ISL_2007483 | Laboratorio de Virología del Hospital de Niños Dr. Ricardo Gutierrez | Área de Secuenciación del Laboratorio de Virología del Hospital de Niños Dr. Ricardo Gutierrez on behalf of 'Proyecto Argentino Interinstitucional de genomica de SARS-CoV-2' (PAIS Consortium) | Alexay, S; Thomas, G; Medina, C; Labarta, N; Streitenberger, C; Villegas, E; Barreda Frank, M; Grandis, E; Acevedo, ME; Alvarez Lopez, C; Jacques, O; Mistchenko, A; Nabaes Jodar, M; Goya, S; Lusso, S; Acuña, D; Natale, MI; Valinotto, LE; Viegas, M. |
| EPI_ISL_2007484 | Hospital General de Agudos Dr. Cosme Argerich | Área de Secuenciación del Laboratorio de Virología del Hospital de Niños Dr. Ricardo Gutierrez on behalf of 'Proyecto Argentino Interinstitucional de genomica de SARS-CoV-2' (PAIS Consortium) | Marcia Pozzati, Jésica Galeano, Florencia Rodríguez, Florencia Funez, Andrea Fernández, Karina Polanski; Alexay, S; Nabaes Jodar, M; Acuña, D; Goya, S; Lusso, S; Natale, MI; Valinotto, LE; Viegas, M. |
| EPI_ISL_2007485, EPI_ISL_2007487, | Laboratorio de Virología del Hospital de Niños Dr. Ricardo | Área de Secuenciación del Laboratorio de Virología del | Alexay, S; Thomas, G; Medina, C; Labarta, N; Streitenberger, C; Villegas, E; Barreda Frank, M; Grandis, E; Acevedo, ME; Alvarez Lopez, C; Jacques, O; |

| | | | |
|---|---|---|---|
| EPI_ISL_2007491, EPI_ISL_2007492, EPI_ISL_2007497, EPI_ISL_2007498, EPI_ISL_2007499, EPI_ISL_2007500 | Gutierrez | Hospital de Niños Dr. Ricardo Gutierrez on behalf of 'Proyecto Argentino Interinstitucional de genomica de SARS-CoV-2' (PAIS Consortium) | Mistchenko, A; Nabaes Jodar, M; Goya, S; Lusso, S; Acuña, D; Natale, MI; Valinotto, LE; Viegas, M. |
| EPI_ISL_2007501 | Laboratorio Central, Ministerio de Salud Córdoba | Instituto de Patologia Vegetal (CIAP-INTA) on behalf of 'Proyecto Argentino Interinstitucional de genomica de SARS-CoV-2' (PAIS Consortium) | Fernández, FD; Marquez, N.; Debat, HJ.; Amadio, A; Irazoqui, M; Re, V.; Pisano, M.B.; Castro, G.; Barbas, G. |
| EPI_ISL_2007505 | Hospital General de Agudos Dr. Cosme Argerich | Área de Secuenciación del Laboratorio de Virología del Hospital de Niños Dr. Ricardo Gutierrez on behalf of 'Proyecto Argentino Interinstitucional de genomica de SARS-CoV-2' (PAIS Consortium) | Marcia Pozzati, Jésica Galeano, Florencia Rodríguez, Florencia Funez, Andrea Fernández, Karina Polanski; Alexay, S; Nabaes Jodar, M; Acuña, D; Goya, S; Lusso, S; Natale, MI; Valinotto, LE; Viegas, M. |
| EPI_ISL_2007506, EPI_ISL_2007507, EPI_ISL_2007508, EPI_ISL_2007509, EPI_ISL_2007510, EPI_ISL_2007511, EPI_ISL_2007512, EPI_ISL_2007513 | Laboratorio de Virología del Hospital de Niños Dr. Ricardo Gutierrez | Área de Secuenciación del Laboratorio de Virología del Hospital de Niños Dr. Ricardo Gutierrez on behalf of 'Proyecto Argentino Interinstitucional de genomica de SARS-CoV-2' (PAIS Consortium) | Alexay, S; Thomas, G; Medina, C; Labarta, N; Streitenberger, C; Villegas, E; Barreda Frank, M; Grandis, E; Acevedo, ME; Alvarez Lopez, C; Jacques, O; Mistchenko, A; Nabaes Jodar, M; Goya, S; Lusso, S; Acuña, D; Natale, MI; Valinotto, LE; Viegas, M. |
| EPI_ISL_2007514, EPI_ISL_2007515, EPI_ISL_2007516 | Laboratorio Central, Ministerio de Salud Córdoba | Instituto de Patologia Vegetal (CIAP-INTA) on behalf of 'Proyecto Argentino Interinstitucional de genomica de SARS-CoV-2' (PAIS Consortium) | Fernández, FD; Marquez, N.; Debat, HJ.; Amadio, A; Irazoqui, M; Re, V.; Pisano, M.B.; Castro, G.; Barbas, G. |
| EPI_ISL_2007517, EPI_ISL_2007518 | Laboratorio de Virología del Hospital de Niños Dr. Ricardo Gutierrez | Área de Secuenciación del Laboratorio de Virología del Hospital de Niños Dr. Ricardo Gutierrez on behalf of 'Proyecto Argentino Interinstitucional de genomica de SARS-CoV-2' (PAIS Consortium) | Alexay, S; Thomas, G; Medina, C; Labarta, N; Streitenberger, C; Villegas, E; Barreda Frank, M; Grandis, E; Acevedo, ME; Alvarez Lopez, C; Jacques, O; Mistchenko, A; Nabaes Jodar, M; Goya, S; Lusso, S; Acuña, D; Natale, MI; Valinotto, LE; Viegas, M. |
| EPI_ISL_2007521 | Hospital General de Agudos Dr. Cosme Argerich | Área de Secuenciación del Laboratorio de Virología del Hospital de Niños Dr. Ricardo Gutierrez on behalf of 'Proyecto Argentino Interinstitucional de genomica de SARS-CoV-2' (PAIS Consortium) | Marcia Pozzati, Jésica Galeano, Florencia Rodríguez, Florencia Funez, Andrea Fernández, Karina Polanski; Alexay, S; Nabaes Jodar, M; Acuña, D; Goya, S; Lusso, S; Natale, MI; Valinotto, LE; Viegas, M. |
| EPI_ISL_2007522, EPI_ISL_2007537, EPI_ISL_2007544 | Laboratorio de Virología del Hospital de Niños Dr. Ricardo Gutierrez | Área de Secuenciación del Laboratorio de Virología del Hospital de Niños Dr. Ricardo Gutierrez on behalf of 'Proyecto Argentino Interinstitucional de genomica de SARS-CoV-2' (PAIS Consortium) | Alexay, S; Thomas, G; Medina, C; Labarta, N; Streitenberger, C; Villegas, E; Barreda Frank, M; Grandis, E; Acevedo, ME; Alvarez Lopez, C; Jacques, O; Mistchenko, A; Nabaes Jodar, M; Goya, S; Lusso, S; Acuña, D; Natale, MI; Valinotto, LE; Viegas, M. |
| EPI_ISL_2010570 | Helix/Illumina | Centers for Disease Control and Prevention Division of Viral Diseases, Pathogen Discovery | Dakota Howard, Dhwani Batra, Peter W. Cook, Kara Moser, Adrian Paskey, Jason Caravas, Benjamin Rambo-Martin, Shatavia Morrison, Christopher Gulvick, Scott Sammons, Yvette Unoarumhi, Darlene Wagner, Matthew Schmerer, Eileen de Feo, Jan Antico, Christine Tran, Matthew Tolentino, Shannon Wickline, Kim Gietzen, Brad Sickler, Jingtao Liu, Eric Allen, Phil Febbo, Nicole L. Washington, Simon White, Geraint Levan, Kelly Schiabor Barrett, Elizabeth Cirulli, Alexandre Bolze, Ary Ascencio, Charlotte Rivera-Garcia, Ryan Cho, Jason Nguyen, Sherry Wang, Jimmy Ramirez, Tyler Cassens, Efren Sandoval, Magnus Isaksson, William Lee, David Becker, Marc Laurent, James Lu, Clinton R. Paden, Duncan MacCannell |
| EPI_ISL_2023146 | Yale Clinical Virology Lab | Grubaugh Lab - Yale School of Public Health | Joseph Fauver, Mallery Breban, Isabel Ott, Tara Alpert, Mary Petrone, Anderson Brito, Chantal Vogels, Annie Watkins, Chaney Kalinich, Jessica Rothman, Marie L. Landry, Nathan Grubaugh |
| EPI_ISL_2040651, EPI_ISL_2042190, EPI_ISL_2042280 | Aegis Sciences Corporation | Centers for Disease Control and Prevention Division of Viral Diseases, Pathogen Discovery | Dakota Howard, Dhwani Batra, Peter W. Cook, Kara Moser, Adrian Paskey, Jason Caravas, Benjamin Rambo-Martin, Shatavia Morrison, Christopher Gulvick, Scott Sammons, Yvette Unoarumhi, Darlene Wagner, Matthew Schmerer, Cyndi Clark, Patrick Campbell, Rob Case, Vikramsinha Ghorpade, Holly Houdeshell, Ola Kvalvaag, Dillon Nall, Ethan Sanders, Alec Vest, Shaun Westlund, Matthew Hardison, Clinton R. Paden, Duncan MacCannell |
| EPI_ISL_2083473 | Israel Central Virology laboratory | Israel National Consortium for SARS-CoV-2 sequencing | Neta Zuckerman, Efrat Dahan Bucris, Michal Mandelboim, Dana Bar-Ilan, Oran Erster, Tzvia Mann, Omer Murik, David A. Zeevi, Assaf Rokney, Joseph Jaffe, Eva Nachum, Maya Davidovich Cohen, Ephraim Fass, Gal Zizelski Valenci, Mor Rubinstein, Efrat Rorman, Israel Nissan, Efrat Glick-Saar, Omri Nayshool, Gideon Rechavi, Ella Mendelson, Orna Mor |
| EPI_ISL_2085580 | Israel Central Virology laboratory | Israel National Consortium for SARS-CoV-2 sequencing | Neta Zuckerman, Efrat Dahan Bucris, Michal Mandelboim, Dana Bar-Ilan, Miranda Geva, Netanel Abu, Oran Erster, Efrat Glick-Saar, Omri Nayshool, Gideon Rechavi, Ella Mendelson, Orna Mor |
| EPI_ISL_2089827 | Aegis Sciences Corporation | Centers for Disease Control and Prevention Division of Viral Diseases, Pathogen Discovery | Dakota Howard, Dhwani Batra, Peter W. Cook, Kara Moser, Adrian Paskey, Jason Caravas, Benjamin Rambo-Martin, Shatavia Morrison, Christopher Gulvick, Scott Sammons, Yvette Unoarumhi, Darlene Wagner, Matthew Schmerer, Cyndi Clark, Patrick Campbell, Rob Case, Vikramsinha Ghorpade, Holly Houdeshell, Ola Kvalvaag, Dillon Nall, Ethan Sanders, Alec Vest, Shaun Westlund, Matthew Hardison, Clinton R. Paden, Duncan MacCannell |
| EPI_ISL_2096430 | Broad Institute Clinical Research Sequencing Platform | Infectious Disease Program, Broad Institute of Harvard and MIT | Siddle,K.J., Adams,G., Pearlman,L., Gladden-Young,A., Vicente,G., Blumenstiel,B., DeFelice,M., Lee,M., McGovern,S., Lagerborg,K., Rudy,M., DeRuff,K., Carter,A., Normandin,E., Bauer,M., Reilly,S., Tomkins-Tinch,C., Loreth,C., Chaluvadi,S., Meldrim,J., Granger,B., Lemieux,J.E., Birren,B.W., Sabeti,P.C., Larkin,K., Dodge,S., Lennon,N., Madoff,L., Brown,C., Gallagher,G., Smole,S., Park,D.J., Gabriel,S., and MacInnis,B.L. |
| EPI_ISL_2096846, EPI_ISL_2096856 | Hadassah Medical Center Clinical Virology Laboratory, Hadassah Ein Kerem | Hadassah Hebrew University Viral Sequencing Group, Hadassah Hebrew University Medical Center | Hadar Golan Berman, Esther Oiknine-Djian, Mila Rivkin, Sheera Adar, Dana G. Wolf |
| EPI_ISL_2101894 | LESP Guanajuato | Instituto de Diagnostico y Referencia Epidemiologicos (INDRE) | Claudia Wong-Arambula, Abril Rodriguez-Maldonado, Vanessa Rivero-Arredondo, Ariadna Medina-Benitez, Joaquin Quiroz-Mercado, Sergio Rangel-Guerrero, Natividad Cruz-Ortiz, Tatiana Nunez-Garcia, Gisela Barrera-Badillo, Lucia Hernandez-Rivas, Irma Lopez-Martinez, Ernesto Ramirez-Gonzalez. |
| EPI_ISL_2101899 | LESP Quintana Roo | Instituto de Diagnostico y Referencia Epidemiologicos (INDRE) | Claudia Wong-Arambula, Abril Rodriguez-Maldonado, Vanessa Rivero-Arredondo, Ariadna Medina-Benitez, Joaquin Quiroz-Mercado, Sergio Rangel-Guerrero, Natividad Cruz-Ortiz, Tatiana Nunez-Garcia, Gisela Barrera-Badillo, Lucia Hernandez-Rivas, Irma Lopez-Martinez, Ernesto Ramirez-Gonzalez. |
| EPI_ISL_2102372, EPI_ISL_2102374, EPI_ISL_2102375 | Maryland Genomics, Institute for Genome Sciences, University of Maryland School of Medicine | Maryland Genomics, Institute for Genome Sciences, University of Maryland School of Medicine | Tallon, Luke J; Sadzewicz, Lisa D; Humphrys, Mike; Ott, Sandra; Roussey, Holly; Mehta, Aditya; Vavikolanu, Kranthi; Fraser, Claire M; Ravel, Jacques |
| EPI_ISL_2105822 | Salud Digna | Instituto Nacional de Medicina Genomica | Hidalgo-Miranda A, Cedro-Tanda A, Mendoza-Vargas A, Reyes-Grajeda JP, Abraham Campos-Romero, Moreno-Camacho José Luis, Rodriguez-Gallegos Jorge, Luna-Ruiz Marco, Gonzalez-Barrera D, Rangel-DeLeon D, Munguia-Garza P, Ramirez-Vega O, Escobar-Arrazola, M, Herrera-Montalvo LA. |
| EPI_ISL_2109730 | Sonic - Labor Dr. von Froreich GmbH | Robert Koch Institute | unknown |
| EPI_ISL_2122995 | Labor Dr. Heidrich & Kollegen MVZ GmbH Hamburg | Robert Koch Institute | unknown |
| EPI_ISL_2133176 | Quest Diagnostics Incorporated | Centers for Disease Control and Prevention Division of Viral Diseases, Pathogen Discovery | Dakota Howard, Dhwani Batra, Peter W. Cook, Kara Moser, Adrian Paskey, Jason Caravas, Benjamin Rambo-Martin, Shatavia Morrison, Christopher Gulvick, Scott Sammons, Yvette Unoarumhi, Darlene Wagner, Matthew Schmerer, S. H. Rosenthal, A. Gerasimova, R. M. Kagan, B. Anderson, M. Hua, Y. Liu, L.E. Bernstein, K.E. Livingston, A. Perez, I. A. Shlyakhter, R. V. Rolando, R. Owen, P. Tanpaiboon, F. Lacbawan, Clinton R. Paden, Duncan MacCannell |
| EPI_ISL_2133399, EPI_ISL_2133401 | Servicio Virosis Respiratorias-Departamento Virología-INEI | Instituto Nacional Enfermedades Infecciosas C.G.Malbran | Baumeister E., Avaro M., Benedetti E., Russo M., Dattero ME, Pontoriero A., Cisterna D., Molina V., Perandones C., Tuduri E., Lorenzo F., Poklepovich T., Campos J. |
| EPI_ISL_2133705 | Quest Diagnostics Incorporated | Centers for Disease Control and Prevention Division of Viral Diseases, Pathogen Discovery | Dakota Howard, Dhwani Batra, Peter W. Cook, Kara Moser, Adrian Paskey, Jason Caravas, Benjamin Rambo-Martin, Shatavia Morrison, Christopher Gulvick, Scott Sammons, Yvette Unoarumhi, Darlene Wagner, Matthew Schmerer, S. H. Rosenthal, A. Gerasimova, R. M. Kagan, B. Anderson, M. Hua, Y. Liu, L.E. Bernstein, K.E. Livingston, A. Perez, I. A. Shlyakhter, R. V. Rolando, R. Owen, P. Tanpaiboon, F. Lacbawan, Clinton R. Paden, Duncan MacCannell |

EPI_ISL_2135137, EPI_ISL_2135150, EPI_ISL_2135255, EPI_ISL_2135326, EPI_ISL_2135327, EPI_ISL_2135336, EPI_ISL_2135337, EPI_ISL_2135338, EPI_ISL_2135342, EPI_ISL_2135697, EPI_ISL_2135698, EPI_ISL_2135699, EPI_ISL_2135722, EPI_ISL_2135723, EPI_ISL_2135725, EPI_ISL_2136003, EPI_ISL_2136010,

| | | | |
|---|---|---|---|
| EPI_ISL_2136023, EPI_ISL_2136027, EPI_ISL_2136028, EPI_ISL_2136036, EPI_ISL_2136044, EPI_ISL_2136046, EPI_ISL_2136049, EPI_ISL_2136078, EPI_ISL_2136079, EPI_ISL_2136080, EPI_ISL_2136081, EPI_ISL_2136083, EPI_ISL_2136084, EPI_ISL_2136085, EPI_ISL_2136086, EPI_ISL_2136087, EPI_ISL_2136088, EPI_ISL_2136089, EPI_ISL_2136091, EPI_ISL_2136093, EPI_ISL_2136095, EPI_ISL_2136098, EPI_ISL_2136140, EPI_ISL_2136149, EPI_ISL_2136164, EPI_ISL_2136178, EPI_ISL_2136179, EPI_ISL_2136180 | | | |
| see above | Servicio Virosis Respiratorias-Departamento Virología-INEI | Instituto Nacional Enfermedades Infecciosas C.G.Malbran | Baumeister E., Avaro M., Benedetti E., Russo M., Dattero ME, Pontoriero A., Cisterna D., Molina V., Perandones C., Tuduri E., Lorenzo F., Poklepovich T., Campos J. |
| EPI_ISL_2139974 | Illinois Department of Public Health | Illinois Department of Public Health - Chicago Lab | Vineet K. Dhiman, Ira Heimler, Joel Price |
| EPI_ISL_2140053, EPI_ISL_2140056, EPI_ISL_2140064, EPI_ISL_2140068, EPI_ISL_2140070, EPI_ISL_2140074, EPI_ISL_2140104, EPI_ISL_2140105, EPI_ISL_2140110, EPI_ISL_2140132 | Servicio Virosis Respiratorias-Departamento Virología-INEI | Instituto Nacional Enfermedades Infecciosas C.G.Malbran | Baumeister E., Avaro M., Benedetti E., Russo M., Dattero ME, Pontoriero A., Cisterna D., Molina V., Perandones C., Tuduri E., Lorenzo F., Poklepovich T., Campos J. |
| EPI_ISL_2148039, EPI_ISL_2148642, EPI_ISL_2149693, EPI_ISL_2149704, EPI_ISL_2149794, EPI_ISL_2150432, EPI_ISL_2150525, EPI_ISL_2150807 | Aegis Sciences Corporation | Centers for Disease Control and Prevention Division of Viral Diseases, Pathogen Discovery | Dakota Howard, Dhwani Batra, Peter W. Cook, Kara Moser, Adrian Paskey, Jason Caravas, Benjamin Rambo-Martin, Shatavia Morrison, Christopher Gulvick, Scott Sammons, Yvette Unoarumhi, Darlene Wagner, Matthew Schmerer, Cyndi Clark, Patrick Campbell, Rob Case, Vikramsinha Ghorpade, Holly Houdeshell, Ola Kvalvaag, Dillon Nall, Ethan Sanders, Alec Vest, Shaun Westlund, Matthew Hardison, Clinton R. Paden, Duncan MacCannell |
| EPI_ISL_2158694, EPI_ISL_2158696, EPI_ISL_2158699, EPI_ISL_2158711, EPI_ISL_2158719, EPI_ISL_2158725, EPI_ISL_2158726, EPI_ISL_2158731, EPI_ISL_2158733, EPI_ISL_2158736, EPI_ISL_2158743, EPI_ISL_2158749, EPI_ISL_2158751, EPI_ISL_2158775, EPI_ISL_2158779, EPI_ISL_2158780, EPI_ISL_2158784, EPI_ISL_2158786, EPI_ISL_2158791, EPI_ISL_2158804, EPI_ISL_2158806, EPI_ISL_2158819, EPI_ISL_2158826, EPI_ISL_2158830, EPI_ISL_2158837, EPI_ISL_2158842, EPI_ISL_2158848 | | | |
| see above | Servicio Virosis Respiratorias-Departamento Virología-INEI | Instituto Nacional Enfermedades Infecciosas C.G.Malbran | Baumeister E., Avaro M., Benedetti E., Russo M., Dattero ME, Pontoriero A., Cisterna D., Molina V., Perandones C., Tuduri E., Lorenzo F., Poklepovich T., Campos J. |
| EPI_ISL_2159924, EPI_ISL_2159930, EPI_ISL_2159981, EPI_ISL_2160142 | Helix/Illumina | Centers for Disease Control and Prevention Division of Viral Diseases, Pathogen Discovery | Dakota Howard, Dhwani Batra, Peter W. Cook, Kara Moser, Adrian Paskey, Jason Caravas, Benjamin Rambo-Martin, Shatavia Morrison, Christopher Gulvick, Scott Sammons, Yvette Unoarumhi, Darlene Wagner, Matthew Schmerer, Eileen de Feo, Jan Antico, Christine Tran, Matthew Tolentino, Shannon Wickline, Kim Gietzen, Brad Sickler, Jingtao Liu, Eric Allen, Phil Febbo, Nicole L. Washington, Simon White, Geraint Levan, Kelly Schiabor Barrett, Elizabeth Cirulli, Alexandre Bolze, Ary Ascencio, Charlotte Rivera-Garcia, Ryan Cho, Jason Nguyen, Sherry Wang, Jimmy Ramirez, Tyler Cassens, Efren Sandoval, Magnus Isaksson, William Lee, David Becker, Marc Laurent, James Lu, Clinton R. Paden, Duncan MacCannell |
| EPI_ISL_2161453 | Maryland Genomics, Institute for Genome Sciences, University of Maryland School of Medicine | Maryland Genomics, Institute for Genome Sciences, University of Maryland School of Medicine | Tallon, Luke J; Sadzewicz, Lisa D; Humphrys, Mike; Ott, Sandra; Roussey, Holly; Mehta, Aditya; Vavikolanu, Kranthi; Fraser, Claire M; Ravel, Jacques |
| EPI_ISL_2180667, EPI_ISL_2181126 | Aegis Sciences Corporation | Centers for Disease Control and Prevention Division of Viral Diseases, Pathogen Discovery | Dakota Howard, Dhwani Batra, Peter W. Cook, Kara Moser, Adrian Paskey, Jason Caravas, Benjamin Rambo-Martin, Shatavia Morrison, Christopher Gulvick, Scott Sammons, Yvette Unoarumhi, Darlene Wagner, Matthew Schmerer, Cyndi Clark, Patrick Campbell, Rob Case, Vikramsinha Ghorpade, Holly Houdeshell, Ola Kvalvaag, Dillon Nall, Ethan Sanders, Alec Vest, Shaun Westlund, Matthew Hardison, Clinton R. Paden, Duncan MacCannell |
| EPI_ISL_2182966, EPI_ISL_2183218, EPI_ISL_2183609, EPI_ISL_2183795 | Israel Central Virology laboratory | Israel National Consortium for SARS-CoV-2 sequencing | Neta Zuckerman, Efrat Dahan Bucris, Michal Mandelboim, Dana Bar-Ilan, Miranda Geva, Netanel Abu, Oran Erster, Efrat Glick-Saar, Omri Nayshool, Gideon Rechavi, Ella Mendelson, Orna Mor |
| EPI_ISL_2186247, EPI_ISL_2187561 | Aegis Sciences Corporation | Centers for Disease Control and Prevention Division of Viral Diseases, Pathogen Discovery | Dakota Howard, Dhwani Batra, Peter W. Cook, Kara Moser, Adrian Paskey, Jason Caravas, Benjamin Rambo-Martin, Shatavia Morrison, Christopher Gulvick, Scott Sammons, Yvette Unoarumhi, Darlene Wagner, Matthew Schmerer, Cyndi Clark, Patrick Campbell, Rob Case, Vikramsinha Ghorpade, Holly Houdeshell, Ola Kvalvaag, Dillon Nall, Ethan Sanders, Alec Vest, Shaun Westlund, Matthew Hardison, Clinton R. Paden, Duncan MacCannell |
| EPI_ISL_2192957 | Broad Institute Clinical Research Sequencing Platform | Infectious Disease Program, Broad Institute of Harvard and MIT | Siddle,K.J., Adams,G., Pearlman,L., Gladden-Young,A., Vicente,G., Blumenstiel,B., DeFelice,M., Lee,M., McGovern,S., Lagerborg,K., Rudy,M., DeRuff,K., Carter,A., Normandin,E., Bauer,M., Reilly,S., Tomkins-Tinch,C., Loreth,C., Chaluvadi,S., Meldrim,J., Granger,B., Lemieux,J.E., Birren,B.W., Sabeti,P.C., Larkin,K., Dodge,S., Lennon,N., Madoff,L., Brown,C., Gallagher,G., Smole,S., Park,D.J., Gabriel,S., and MacInnis,B.L. |
| EPI_ISL_2201766 | Aegis Sciences Corporation | Centers for Disease Control and Prevention Division of Viral Diseases, Pathogen Discovery | Dakota Howard, Dhwani Batra, Peter W. Cook, Kara Moser, Adrian Paskey, Jason Caravas, Benjamin Rambo-Martin, Shatavia Morrison, Christopher Gulvick, Scott Sammons, Yvette Unoarumhi, Darlene Wagner, Matthew Schmerer, Cyndi Clark, Patrick Campbell, Rob Case, Vikramsinha Ghorpade, Holly Houdeshell, Ola Kvalvaag, Dillon Nall, Ethan Sanders, Alec Vest, Shaun Westlund, Matthew Hardison, Clinton R. Paden, Duncan MacCannell |
| EPI_ISL_2211703 | Viollier AG | Department of Biosystems Science and Engineering, ETH Zürich | Christian Beisel, Sarah Nadeau, Chaoran Chen, Ivan Topolsky, Philipp Jablonski, Lara Fuhrmann, David Dreifuss, Katharina Jahn, Rebecca Denes, Mirjam Feldkamp, Ina Nissen, Natascha Santacroce, Elodie Burcklen, Christiane Beckmann, Maurice Redondo, Olivier Kobel, Christoph Noppen, Sophie Seidel, Noemie Santamaria de Souza, Niko Beerenwinkel, Tanja Stadler |
| EPI_ISL_2230870 | Salud Digna | Instituto Nacional de Medicina Genomica | Hidalgo-Miranda A, Cedro-Tanda A, Mendoza-Vargas A, Reyes-Grajeda JP, Abraham Campos-Romero, Moreno-Camacho José Luis, Rodriguez-Gallegos Jorge, Luna-Ruiz Marco, Gonzalez-Barrera D, Rangel-DeLeon D, Munguia-Garza P, Ramirez-Vega O, Escobar-Arrazola, M, Herrera-Montalvo LA. |
| EPI_ISL_2235272, EPI_ISL_2235283, EPI_ISL_2235291 | Department of Microbiology, AHEPA University Hospital | Institute of Applied Biosciences, Centre for Research and Technology Hellas | Anastasia Chatzidimitriou et al. |
| EPI_ISL_2238524 | Lighthouse Lab in Glasgow | Wellcome Sanger Institute for the COVID-19 Genomics UK (COG-UK) Consortium | Harper VanSteenhouse, Yumi Kasai, David Gray, Carol Clugston, Anna Dominiczak and Alex Alderton, Roberto Amato, Jeffrey Barrett, Sonia Goncalves, Ewan Harrison, David K. Jackson, Ian Johnston, Dominic Kwiatkowski, Cordelia Langford, John Sillitoe on behalf of the Wellcome Sanger Institute COVID-19 Surveillance Team |
| EPI_ISL_2239796 | Northumbria University / South Tees Hospitals NHS Foundation Trust / North Cumbria Integrated Care NHS Foundation Trust / North Tees and Hartlepool NHS Foundation Trust / Newcastle Hospitals NHS Foundation Trust | COVID-19 Genomics UK (COG-UK) Consortium | Darren L Smith,Andrew Nelson,Matthew Bashton,Greg R Young,Joshua Loh,John Allan,Mohammad A Tariq,Giles S Holt,Gary Black,Wen C Yew,Lynn Dover,Paul Baker,Steve Liggett,Sarah Essex,Jane Greenaway,Debra Padgett,Clive Graham,Garren Scott,Edward Barton,Emma Swindells,Brendan Payne,Jennifer Collins,Yusri Taha,Gary Eltringham |
| EPI_ISL_2240466 | Oxford Viromics, NDM, University of Oxford; Oxford University Hospitals; Basingstoke and North Hampshire Hospital | COVID-19 Genomics UK (COG-UK) Consortium | Tanya Golubchik, David Bonsall, George Macintyre, Amy Trebes, Mariateresa de Cesare, Catrin Moore, Alex Mobbs, Anita Justice, Robert Shaw, Monique Andersson, Timothy Peto, Emma Wise, Nathan Moore, Jessica Lynch, Nick Cortes, Matilde Mori, Stephen Kidd, David Buck, John Todd, Christophe Fraser |
| EPI_ISL_2241736, EPI_ISL_2241972, EPI_ISL_2242219, EPI_ISL_2242331, EPI_ISL_2242452 | Aegis Sciences Corporation | Centers for Disease Control and Prevention Division of Viral Diseases, Pathogen Discovery | Dakota Howard, Dhwani Batra, Peter W. Cook, Kara Moser, Adrian Paskey, Jason Caravas, Benjamin Rambo-Martin, Shatavia Morrison, Christopher Gulvick, Scott Sammons, Yvette Unoarumhi, Darlene Wagner, Matthew Schmerer, Cyndi Clark, Patrick Campbell, Rob Case, Vikramsinha Ghorpade, Holly Houdeshell, Ola Kvalvaag, Dillon Nall, Ethan Sanders, Alec Vest, Shaun Westlund, Matthew Hardison, Clinton R. Paden, Duncan MacCannell |
| EPI_ISL_2249836, EPI_ISL_2250151 | Laboratory Medicine | Department of Laboratory Medicine, Lin-Kou Chang Gung Memorial Hospital, Taoyuan, Taiwan | Kuo-Chien Tsao, Yu-Nong Gong, Shu-Li Yang, Yi-Chun Liu, Chung-Guei Huang, Mei-Jen Hsiao, Po-Wei Huang, Cheng-Ta Yang, Cheng-Hsun Chiu, Peng-Nien Huang, Kuo-Ming Lee, Guang-Wu Chen, Shin-Ru Shih |
| EPI_ISL_2258492 | Department of Virology and Immunology, University of Helsinki and Helsinki University Hospital, Huslab Finland | Department of Virology, Faculty of Medicine, University of Helsinki, Helsinki, Finland | Teemu Smura, Ravi Kant, Phuoc Truong, Hussein Alburkat, Hannimari Kallio-Kokko, Jenni Virtanen, Maija Suvanto, Essi Korhonen, Sari Hannula, Harri Kangas, Hanna Liimatainen, Satu Kurkela, Hanna Jarva, Maija Lappalainen, Pekka Ellonen, Olli Vapalahti |
| EPI_ISL_2260334 | Labor 28 MVZ GmbH | Robert Koch Institute | unknown |
| EPI_ISL_2268394, EPI_ISL_2268404, EPI_ISL_2268427 | Quest Diagnostics Incorporated | Centers for Disease Control and Prevention Division of Viral Diseases, Pathogen Discovery | Dakota Howard, Dhwani Batra, Peter W. Cook, Kara Moser, Adrian Paskey, Jason Caravas, Benjamin Rambo-Martin, Shatavia Morrison, Christopher Gulvick, Scott Sammons, Yvette Unoarumhi, Darlene Wagner, Matthew Schmerer, S. H. Rosenthal, A. Gerasimova, R. M. Kagan, B. Anderson, M. Hua, Y. Liu, L.E. Bernstein, K.E. Livingston, A. Perez, I. A. Shlyakhter, R. V. Rolando, R. Owen, P. Tanpaiboon, F. Lacbawan, Clinton R. Paden, Duncan MacCannell |
| EPI_ISL_2270925 | SIESP DIPARTIMENTO DI PREVENZIONE TERAMO | Istituto Zooprofilattico Sperimentale dell'Abruzzo e Molise "G. Caporale" | Lorusso A, Marcacci M, Di Domenico M, Ancora M, Curini V, Di Lollo Valeria, Mangone I, Rinaldi A, Delli Compagni E, Scialabba S, Caporale M, Di Pasquale A, Cammà C, Puglia I, Calistri P, Savini G |
| EPI_ISL_2272583 | GA Department of Public Health | GA Department of Public Health | Stacy Reeves, Jonathan Edwards, Cynthia Dixey, Tonia Parrott, Aliyah Fields, Taylor Smith |
| EPI_ISL_2272976 | AREA DE SALUD SAN JUAN-SAN DIEGO-CONCEPCION 2 | Inciensa, Instituto Costarricense de Investigación y Enseñanza en Nutrición y Salud | Francisco Duarte, Hebleen Porras, Claudio Soto-Garita, Estela Cordero, Adriana Godínez, Melany Calderón, José Luis Vargas, Mariela Gutiérrez, Joselyn Prado & Mariel López |
| EPI_ISL_2272985 | HOSPITAL CIUDAD NEILY | Inciensa, Instituto Costarricense de Investigación y | Francisco Duarte, Hebleen Porras, Claudio Soto-Garita, Estela Cordero, Adriana Godínez, Melany Calderón, José Luis Vargas, Mariela Gutiérrez, Joselyn |

| | | | |
|---|---|---|---|
| | | Enseñanza en Nutrición y Salud | Prado & Raúl Zeledón-Mayorga |
| EPI_ISL_2277760, EPI_ISL_2277780 | Florida Bureau of Public Health Laboratories | Florida Bureau of Public Health Laboratories | Sarah Schmedes, Jason Blanton |
| EPI_ISL_2279168 | Wisconsin State Laboratory of Hygiene Communicable Disease Division | Wisconsin State Laboratory of Hygiene Communicable Disease Division | Abigail C. Shockey, Alicia J. Mooney, Erika M. Hanson, Tonya Danz, Richard Griesser, Sara Wagner, Kelsey R. Florek |
| EPI_ISL_2280581 | Aegis Sciences Corporation | Centers for Disease Control and Prevention Division of Viral Diseases, Pathogen Discovery | Dakota Howard, Dhwani Batra, Peter W. Cook, Kara Moser, Adrian Paskey, Jason Caravas, Benjamin Rambo-Martin, Shatavia Morrison, Christopher Gulvick, Scott Sammons, Yvette Unoarumhi, Darlene Wagner, Matthew Schmerer, Cyndi Clark, Patrick Campbell, Rob Case, Vikramsinha Ghorpade, Holly Houdeshell, Ola Kvalvaag, Dillon Nall, Ethan Sanders, Alec Vest, Shaun Westlund, Matthew Hardison, Clinton R. Paden, Duncan MacCannell |
| EPI_ISL_2283205 | Weill Cornell Medicine | New York Genome Center | Michael Zody, Andre Corvelo, Dayna M. Oschwald, Samantha Fennessey, Tom Maniatis, Melissa Cushing, Olivier Elemento, Margaret Elizabeth Ross, Chris Mason, Priya Velu, Hanna Rennert, Arryn Craney, Lars F Westblade |
| EPI_ISL_2283558 | Infinity Biologix | Centers for Disease Control and Prevention Division of Viral Diseases, Pathogen Discovery | Dakota Howard, Dhwani Batra, Peter W. Cook, Kara Moser, Adrian Paskey, Jason Caravas, Benjamin Rambo-Martin, Shatavia Morrison, Christopher Gulvick, Scott Sammons, Yvette Unoarumhi, Darlene Wagner, Matthew Schmerer, Christian Bixby, Yihe Wang, Jonathan Schultz, Chirayu Goswami, Russ Hager, Robin Grimwood, Clinton R. Paden, Duncan MacCannell |
| EPI_ISL_2283696 | LESP Michoacan | Instituto de Diagnostico y Referencia Epidemiologicos (INDRE) | Claudia Wong-Arambula, Abril Rodriguez-Maldonado, Vanessa Rivero-Arredondo, Ariadna Medina-Benitez, Joaquin Quiroz-Mercado, Sergio Rangel-Guerrero, Natividad Cruz-Ortiz, Tatiana Nunez-Garcia, Gisela Barrera-Badillo, Lucia Hernandez-Rivas, Irma Lopez-Martinez, Ernesto Ramirez-Gonzalez. |
| EPI_ISL_2293455, EPI_ISL_2293463 | Labo Analyses Med | National Reference Center for Viruses of Respiratory Infections, Institut Pasteur, Paris | Marion Barbet, Sylvie Behillil, Méline Bizard, Angela Brisebarre, Camille Capel, Vincent Enouf, Louise Lefrançois, Frédéric Lemoine, Christophe Malabat, Corinne Maufrais, Etienne Simon-Lorière, Maud Vanpeene, Sylvie Van der Werf ,Philippe Miara |
| EPI_ISL_2295590 | LESP Guanajuato | Instituto de Diagnostico y Referencia Epidemiologicos (INDRE) | Claudia Wong-Arambula, Abril Rodriguez-Maldonado, Vanessa Rivero-Arredondo, Ariadna Medina-Benitez, Joaquin Quiroz-Mercado, Sergio Rangel-Guerrero, Natividad Cruz-Ortiz, Tatiana Nunez-Garcia, Gisela Barrera-Badillo, Lucia Hernandez-Rivas, Irma Lopez-Martinez, Ernesto Ramirez-Gonzalez. |
| EPI_ISL_2296385 | Yale Clinical Virology Lab | Grubaugh Lab - Yale School of Public Health | Joseph Fauver, Mallery Breban, Isabel Ott, Tara Alpert, Mary Petrone, Anderson Brito, Chantal Vogels, Annie Watkins, Chaney Kalinich, Jessica Rothman, Marie L. Landry, Nathan Grubaugh |
| EPI_ISL_2301695 | Laboratory of Hygiene and Epidemiology, Department of Medicine, University of Thessaly | Greek Genome Center, Biomedical Research Foundation of the Academy of Athens (BRFAA) | Emmanouil Athanasiadis, Giannis Vatsellas, Theodoros Loupis, Katerina Zoi, Christos Hadjichristodoulou, Dimitrios Thanos |
| EPI_ISL_2308592 | OSPEDALE CIVILE SANT'OMERO - PRONTO SOCCORSO | Istituto Zooprofilattico Sperimentale dell'Abruzzo e Molise "G. Caporale" | Lorusso A, Marcacci M, Di Domenico M, Ancora M, Curini V, Di Lollo Valeria, Mangone I, Rinaldi A, Delli Compagni E, Scialabba S, Caporale M, Di Pasquale A, Cammà C, Puglia I, Calistri P, Savini G |
| EPI_ISL_2319317 | INDRE | Instituto Nacional de Medicina Genomica | Hidalgo-Miranda A, Mendoza-Vargas A, Reyes-Grajeda JP, Cedro-Tanda A, Gisela Barrera-Badillo, Irma Lopez-Martinez, Jose Ernesto Ramirez González, Gonzalez-Barrera D, Rangel-DeLeon D, Munguia-Garza P, Garcia-Cardenas FJ, Gonzalez-Woge MA, Herrera-Montalvo LA. |
| EPI_ISL_2319475, EPI_ISL_2319486, EPI_ISL_2319547, EPI_ISL_2319550 | Maryland Genomics, Institute for Genome Sciences, University of Maryland School of Medicine | Maryland Genomics, Institute for Genome Sciences, University of Maryland School of Medicine | Tallon, Luke J; Sadzewicz, Lisa D; Humphrys, Mike; Ott, Sandra; Roussey, Holly; Mehta, Aditya; Vavikolanu, Kranthi; Fraser, Claire M; Ravel, Jacques |
| EPI_ISL_2319894 | Reditus Laboratories | Reditus Laboratories | Joshua J. Geltz, Ph.D., Robert M. Sgambelluri, Ph.D., Rex Dyer, Ph.D., Cassy Philips, M.S., Alexa Eichelberger, M.S. |
| EPI_ISL_2320418 | Helix/Illumina | Centers for Disease Control and Prevention Division of Viral Diseases, Pathogen Discovery | Dakota Howard, Dhwani Batra, Peter W. Cook, Kara Moser, Adrian Paskey, Jason Caravas, Benjamin Rambo-Martin, Shatavia Morrison, Christopher Gulvick, Scott Sammons, Yvette Unoarumhi, Darlene Wagner, Matthew Schmerer, Eileen de Feo, Jan Antico, Christine Tran, Matthew Tolentino, Shannon Wickline, Kim Gietzen, Brad Sickler, Jingtao Liu, Eric Allen, Phil Febbo, Nicole L. Washington, Simon White, Geraint Levan, Kelly Schiabor Barrett, Elizabeth Cirulli, Alexandre Bolze, Ary Ascencio, Charlotte Rivera-Garcia, Ryan Cho, Jason Nguyen, Sherry Wang, Jimmy Ramirez, Tyler Cassens, Efren Sandoval, Magnus Isaksson, William Lee, David Becker, Marc Laurent, James Lu, Clinton R. Paden, Duncan MacCannell |
| EPI_ISL_2331480, EPI_ISL_2331509, EPI_ISL_2331622, EPI_ISL_2331624, EPI_ISL_2331625, EPI_ISL_2331644 | Johns Hopkins Hospital Department of Pathology | Johns Hopkins Hospital Department of Pathology | C. Paul Morris, Chun Huai Luo, Adannaya Amadi, Matthew Schwartz, Nicholas Gallagher, Heba H. Mostafa |
| EPI_ISL_2331853 | Platform BIS UZA/UAntwerpen | Labo Klinische Biologie, UZA | Marie Le Mercier, Jasmine Coppens, Basil Britto Xavier, Christine Lammens, Veerle Matheeussen, Herman Goossens |
| EPI_ISL_2332660 | Maple Grove Hospital | Minnesota Department of Health, Public Health Laboratory | Alexandra Lorentz, Jacob Garfin, Matt Plumb, and Xiong Wang |
| EPI_ISL_2333996 | BIOMNIS EUROFINS IVRY | Department of Virology, Henri Mondor University Hospital, Assistance Publique Hôpitaux de Paris, Université Paris-Est Créteil, INSERM U955 | Christophe Rodriguez, Slim Fourati, Vanessa Demontant, Guillaume Gricourt, Melissa N'Debi, Alexandre Soulier, Elisabeth Trawinski, Jean-Michel Pawlotsky |
| EPI_ISL_2336204 | Washington State Department of Health Public Health Laboratories | Washington State Department of Health Public Health Laboratories | Drew MacKellar, Philip Dykema, Denny Russell, Joenice Gonzalez, Hannah Gray, Geoff Melly, Vanessa De Los Santos, Darren Lucas, JohnAric Peterson, Avi Singh, Rebecca Cao |
| EPI_ISL_2339310 | Oregon State Public Health Laboratory | Oregon State Public Health Laboratory | Rafia Razzaque, Eugene Yeboah, Vanda Makris, Laura Tsaknaridis, John Fontana and Shane Sevey |
| EPI_ISL_2341367, EPI_ISL_2341390, EPI_ISL_2341407 | The Ohio State University Applied Microbiology Services Laboratory | The Ohio State University Applied Microbiology Services Laboratory | Seth A. Faith PhD |
| EPI_ISL_2341501 | Humboldt County Public Health Laboratory | Chan-Zuckerberg Biohub | CZB Cliahub Consortium |
| EPI_ISL_2341515 | Contra Costa County Public Health Lab | Chan-Zuckerberg Biohub | CZB Cliahub Consortium |
| EPI_ISL_2341644 | CA DPH Viral and Rickettsial Disease Laboratory | Chan-Zuckerberg Biohub | CZB Cliahub Consortium |
| EPI_ISL_2343034 | University Hospitals of Geneva, Laboratory of Virology | HUG, Laboratory of Virology and the Health2030 Genome Center | Samuel Cordey, Ana Rita Goncalves, Laurent Kaiser, Lorenzo Cerutti, Henri Pegeot, Melyssa Elies, Deborah Penet, Keith Harshman, Ioannis Xenarios, Emmanouil Dermitzakis |
| EPI_ISL_2343466, EPI_ISL_2343697, EPI_ISL_2343792 | Greek Genome Center, Biomedical Research Foundation of the Academy of Athens (BRFAA) | Greek Genome Center, Biomedical Research Foundation of the Academy of Athens (BRFAA) | Emmanouil Athanasiadis, Giannis Vatsellas, Theodoros Loupis, Katerina Zoi, Dimitrios Thanos |
| EPI_ISL_2346103 | Oregon State Public Health Laboratory | Oregon State Public Health Laboratory | Rafia Razzaque, Eugene Yeboah, Vanda Makris, Laura Tsaknaridis, John Fontana and Shane Sevey |
| EPI_ISL_2346127 | Emory Molecular Diagnostics Laboratory, Emory Healthcare | Piantadosi Lab, Emory Department of Pathology | Ahmed Babiker, Anne Piantadosi |
| EPI_ISL_2347612 | Health Services Laboratories | Wellcome Sanger Institute for the COVID-19 Genomics UK (COG-UK) Consortium | Health Services Laboratories and Alex Alderton, Roberto Amato, Jeffrey Barrett, Sonia Goncalves, Ewan Harrison, David K. Jackson, Ian Johnston, Dominic Kwiatkowski, Cordelia Langford, John Sillitoe on behalf of the Wellcome Sanger Institute COVID-19 Surveillance Team |
| EPI_ISL_2347863 | Lighthouse Laboratory Plymouth | Wellcome Sanger Institute for the COVID-19 Genomics UK (COG-UK) Consortium | Lighthouse Laboratory Plymouth and Alex Alderton, Roberto Amato, Jeffrey Barrett, Sonia Goncalves, Ewan Harrison, David K. Jackson, Ian Johnston, Dominic Kwiatkowski, Cordelia Langford, John Sillitoe on behalf of the Wellcome Sanger Institute COVID-19 Surveillance Team |
| EPI_ISL_2348326 | Lighthouse Lab in Alderley Park | Wellcome Sanger Institute for the COVID-19 Genomics UK (COG-UK) Consortium | Jacquelyn Wynn, Mairead Hyland, The Lighthouse Lab in Alderley Park and Alex Alderton, Roberto Amato, Jeffrey Barrett, Sonia Goncalves, Ewan Harrison, David K. Jackson, Ian Johnston, Dominic Kwiatkowski, Cordelia Langford, John Sillitoe on behalf of the Wellcome Sanger Institute COVID-19 Surveillance Team |
| EPI_ISL_2348722, EPI_ISL_2348723 | Gyncentrum | Gyncentrum | Emilia Morawiec, Pawel Czerwinski, Adam Pudelko, Aleksandra Skubis-Sikora, Magdalena Samul, Agnieszka Polak, Celina Kruszniewska-Rajs, Tomasz Wasik, Robert Wojtyczka, Jolanta Bartosiewicz- Wasik, Maria Maklasinska-Majdanik |
| EPI_ISL_2349657 | Pandemic Response Lab - NYC | Pandemic Response Lab, R&D | Henry Lee, Michael Hammerling, Melissa Hopkins, Cybill del Castillo, Shinyoung Clair Kang, William Ward, Pradeep Bugga, Sol Rey, Dylan Law, Katharine Nelson, Haiping Hao, Jon Laurent |
| EPI_ISL_2350025, EPI_ISL_2350065 | Salud Digna | Instituto Nacional de Medicina Genomica | Hidalgo-Miranda A, Cedro-Tanda A, Mendoza-Vargas A, Reyes-Grajeda JP, Abraham Campos-Romero, Moreno-Camacho José Luis, Rodriguez-Gallegos |

| | | | |
|---|---|---|---|
| | | | Jorge, Luna-Ruiz Marco, Gonzalez-Barrera D, Rangel-DeLeon D, Munguia-Garza P, Ramirez-Vega O, Escobar-Arrazola, M, Herrera-Montalvo LA. |
| EPI_ISL_2353310 | Lighthouse Lab in Milton Keynes | Wellcome Sanger Institute for the COVID-19 Genomics UK (COG-UK) Consortium | The Lighthouse Lab in Milton Keynes and Alex Alderton, Roberto Amato, Jeffrey Barrett, Sonia Goncalves, Ewan Harrison, David K. Jackson, Ian Johnston, Dominic Kwiatkowski, Cordelia Langford, John Sillitoe on behalf of the Wellcome Sanger Institute COVID-19 Surveillance Team |
| EPI_ISL_2353315 | Lighthouse Laboratory Plymouth | Wellcome Sanger Institute for the COVID-19 Genomics UK (COG-UK) Consortium | Lighthouse Laboratory Plymouth and Alex Alderton, Roberto Amato, Jeffrey Barrett, Sonia Goncalves, Ewan Harrison, David K. Jackson, Ian Johnston, Dominic Kwiatkowski, Cordelia Langford, John Sillitoe on behalf of the Wellcome Sanger Institute COVID-19 Surveillance Team |
| EPI_ISL_2353404 | Randox Laboratories | Wellcome Sanger Institute for the COVID-19 Genomics UK (COG-UK) Consortium | Randox Laboratories and Alex Alderton, Roberto Amato, Jeffrey Barrett, Sonia Goncalves, Ewan Harrison, David K. Jackson, Ian Johnston, Dominic Kwiatkowski, Cordelia Langford, John Sillitoe on behalf of the Wellcome Sanger Institute COVID-19 Surveillance Team |
| EPI_ISL_2353558 | Lighthouse Laboratory Plymouth | Wellcome Sanger Institute for the COVID-19 Genomics UK (COG-UK) Consortium | Lighthouse Laboratory Plymouth and Alex Alderton, Roberto Amato, Jeffrey Barrett, Sonia Goncalves, Ewan Harrison, David K. Jackson, Ian Johnston, Dominic Kwiatkowski, Cordelia Langford, John Sillitoe on behalf of the Wellcome Sanger Institute COVID-19 Surveillance Team |
| EPI_ISL_2355576, EPI_ISL_2355948, EPI_ISL_2356492 | Northumbria University / South Tees Hospitals NHS Foundation Trust / North Cumbria Integrated Care NHS Foundation Trust / North Tees and Hartlepool NHS Foundation Trust / Newcastle Hospitals NHS Foundation Trust | COVID-19 Genomics UK (COG-UK) Consortium | Darren L Smith,Andrew Nelson,Matthew Bashton,Greg R Young,Joshua Loh,John Allan,Mohammad A Tariq,Giles S Holt,Gary Black,Wen C Yew,Lynn Dover,Paul Baker,Steve Liggett,Sarah Essex,Jane Greenaway,Debra Padgett,Clive Graham,Garren Scott,Edward Barton,Emma Swindells,Brendan Payne,Jennifer Collins,Yusri Taha,Gary Eltringham |
| EPI_ISL_2362010, EPI_ISL_2362022, EPI_ISL_2362032 | Virginia Division of Consolidated Laboratory Services | Virginia Division of Consolidated Laboratory Services | Virginia DCLS |
| EPI_ISL_2362145, EPI_ISL_2362197 | Virology Laboratory, Scientific Department, Army Medical Center | Virology Laboratory, Scientific Department, Army Medical Center | Silvia Fillo, Riccardo De Sanctis, Antonella Fortunato, Anella Monte, Rossella Brandi, Giulia Campoli, Marzia Cavalli, Lucia Nicosia, Anna Anselmo, Vanessa Vera Fain, Francesco Giordani, Giandomenico Cerreto, Filippo Molinari, Giancarlo Petralito, Florigio Lista, Maria Anna Spinelli, Margherita De Santis |
| EPI_ISL_2363530, EPI_ISL_2363549, EPI_ISL_2363551, EPI_ISL_2363552, EPI_ISL_2363571, EPI_ISL_2363574 | Laboratorio Central de la Ciudad de Santa Fe | Grupo de Genómica y Bioinformática del Instituto de Investigación de la Cadena Láctea CONICET-INTA on behalf of 'Proyecto Argentino Interinstitucional de genomica de SARS-CoV-2' (PAIS Consortium) | Eberhardt, MF; Irazoqui, JM; Ojeda, G; Rompato, G; Mugna, V; Pastor, C; Amadio, AF |
| EPI_ISL_443600 | Department of Pathology, University of Cambridge | Wellcome Sanger Institute for the COVID-19 Genomics UK (COG-UK) consortium | Luke W Meredith, M. Estée Török , Myra Hosmillo, William L. Hamilton, Martin D. Curran, Theresa Feltwell, Grant Hall, Anna Yakovleva, Fahad A Khokhar, Charlotte J. Houldcroft, Laura G Caller, Aminu S. Jahun, Sarah L. Caddy, Ian Goodfellow; and Alex Alderton, Roberto Amato, Sonia Goncalves, Ewan Harrison, David K. Jackson, Ian Johnston, Dominic Kwiatkowski, Cordelia Langford, John Sillitoe on behalf of the Wellcome Sanger Institute COVID-19 Surveillance Team (http://www.sanger.ac.uk/covid-team) |
| EPI_ISL_451934 | Research Unit, University Hospital for Infectious Diseases "Dr. Fran Mihaljevi" | Cicin Sain lab, Helmholtz Centre for Infection Research | Zeeshan Chaudhry, Kathrin Eschke, Željka Maak Šafranko, Ivan-Christian Kurolt |
| EPI_ISL_477779 | University of Birmingham | COVID-19 Genomics UK (COG-UK) Consortium | Institute of Microbiology, University of Birmingham: Claire McMurray, Joanne Stockton, Samuel Nicholls, Radoslaw Poplawski, Will Rowe, Josh Quick, Nicholas Loman. University of Birmingham Testing Laboratory: Celina M Whalley, Andrew Bosworth, Charlotte Poxon, Kasun Wanigasooriya, Oliver Pickles, Mike Kidd, Alex Richter, Andrew D Beggs PHE Heartlands Lab: Husam Osman, Andrew Bosworth. Queen Elizabeth Hospital: Anna Casey |
| EPI_ISL_500152 | Liverpool Clinical Laboratories | COVID-19 Genomics UK (COG-UK) Consortium | Sam Haldenby, Anita Lucaci, Steve Paterson, Julian Hiscox, Alistair Darby, M Almsaud, A Alrezaihi, Muhannad Alruwaili, Stuart D Armstrong, Jones Benjamin, Eleanor G Bentley, Anu Chawla, Jordan J Clark, Angela Cowell, Richard Eccles, Isabel García-Dorival, Matthew Gemmell, Alessandro Gerada, PKF Gilmore, Richard Gregory, Ximeng Han, Catherine Hartley, Margaret Hughes, Miren Iturriza-Gomara, James Johnson, L Luu, Jenifer Manson, Charlotte Nelson, Elaine O'Toole, Cassie Olateju, Rebekah Penrice-Randal , Lucille Rainbow, N.P Randle, Trevor Ian Robinson, Parul Sharma, Ghada T Shawli, James P Stewart, Neil Swainston, Ecaterina Vamos, Joanne Watts, Mark Whitehead |
| EPI_ISL_532975 | Lighthouse Lab in Glasgow | Wellcome Sanger Institute for the COVID-19 Genomics UK (COG-UK) consortium | Harper VanSteenhouse, Yumi Kasai, David Gray, Carol Clugston, Anna Dominiczak and Alex Alderton, Roberto Amato, Sonia Goncalves, Ewan Harrison, David K. Jackson, Ian Johnston, Dominic Kwiatkowski, Cordelia Langford, John Sillitoe |
| EPI_ISL_554361, EPI_ISL_554494 | Lighthouse Lab in Alderley Park | Wellcome Sanger Institute for the COVID-19 Genomics UK (COG-UK) consortium | The Lighthouse Lab in Alderley Park and Alex Alderton, Roberto Amato, Sonia Goncalves, Ewan Harrison, David K. Jackson, Ian Johnston, Dominic Kwiatkowski, Cordelia Langford, John Sillitoe on behalf of the Wellcome Sanger Institute COVID-19 Surveillance Team |
| EPI_ISL_569876 | Amedeo di savoia | Crosetto lab, Karolinska Institutet, SciLifeLab | Michele Simonetti, Maria Grazia Milia, Luuk Harbers, Ning Zhang, Anna Sapino, Valeria Ghisetti, Nicola Crosetto |
| EPI_ISL_600093, EPI_ISL_601443 | Lighthouse Lab in Milton Keynes | Wellcome Sanger Institute for the COVID-19 Genomics UK (COG-UK) consortium | The Lighthouse Lab in Milton Keynes and Alex Alderton, Roberto Amato, Sonia Goncalves, Ewan Harrison, David K. Jackson, Ian Johnston, Dominic Kwiatkowski, Cordelia Langford, John Sillitoe on behalf of the Wellcome Sanger Institute COVID-19 Surveillance Team (http://www.sanger.ac.uk/covid-team) |
| EPI_ISL_633642 | Lighthouse Lab in Glasgow | Wellcome Sanger Institute for the COVID-19 Genomics UK (COG-UK) consortium | Harper VanSteenhouse, Yumi Kasai, David Gray, Carol Clugston, Anna Dominiczak and Alex Alderton, Roberto Amato, Sonia Goncalves, Ewan Harrison, David K. Jackson, Ian Johnston, Dominic Kwiatkowski, Cordelia Langford, John Sillitoe on behalf of the Wellcome Sanger Institute COVID-19 Surveillance Team |
| EPI_ISL_634338 | Lighthouse Lab in Milton Keynes | Wellcome Sanger Institute for the COVID-19 Genomics UK (COG-UK) consortium | The Lighthouse Lab in Milton Keynes and Alex Alderton, Roberto Amato, Sonia Goncalves, Ewan Harrison, David K. Jackson, Ian Johnston, Dominic Kwiatkowski, Cordelia Langford, John Sillitoe on behalf of the Wellcome Sanger Institute COVID-19 Surveillance Team |
| EPI_ISL_655523 | Lighthouse Lab in Alderley Park | Wellcome Sanger Institute for the COVID-19 Genomics UK (COG-UK) Consortium | Jacquelyn Wynn, Mairead Hyland, The Lighthouse Lab in Alderley Park and Alex Alderton, Roberto Amato, Sonia Goncalves, Ewan Harrison, David K. Jackson, Ian Johnston, Dominic Kwiatkowski, Cordelia Langford, John Sillitoe on behalf of the Wellcome Sanger Institute COVID-19 Surveillance Team |
| EPI_ISL_658409 | Lighthouse Lab in Glasgow | Wellcome Sanger Institute for the COVID-19 Genomics UK (COG-UK) Consortium | Harper VanSteenhouse, Yumi Kasai, David Gray, Carol Clugston, Anna Dominiczak and Alex Alderton, Roberto Amato, Sonia Goncalves, Ewan Harrison, David K. Jackson, Ian Johnston, Dominic Kwiatkowski, Cordelia Langford, John Sillitoe on behalf of the Wellcome Sanger Institute COVID-19 Surveillance Team |
| EPI_ISL_659106 | Lighthouse Lab in Cambridge | Wellcome Sanger Institute for the COVID-19 Genomics UK (COG-UK) Consortium | Rob Howes, The Lighthouse Lab in Cambridge and Alex Alderton, Roberto Amato, Sonia Goncalves, Ewan Harrison, David K. Jackson, Ian Johnston, Dominic Kwiatkowski, Cordelia Langford, John Sillitoe on behalf of the Wellcome Sanger Institute COVID-19 Surveillance Team |
| EPI_ISL_659955 | Lighthouse Lab in Glasgow | Wellcome Sanger Institute for the COVID-19 Genomics UK (COG-UK) Consortium | Harper VanSteenhouse, Yumi Kasai, David Gray, Carol Clugston, Anna Dominiczak and Alex Alderton, Roberto Amato, Sonia Goncalves, Ewan Harrison, David K. Jackson, Ian Johnston, Dominic Kwiatkowski, Cordelia Langford, John Sillitoe on behalf of the Wellcome Sanger Institute COVID-19 Surveillance Team |
| EPI_ISL_673868 | Lighthouse Lab in Cambridge | Wellcome Sanger Institute for the COVID-19 Genomics UK (COG-UK) Consortium | Rob Howes, The Lighthouse Lab in Cambridge and Alex Alderton, Roberto Amato, Sonia Goncalves, Ewan Harrison, David K. Jackson, Ian Johnston, Dominic Kwiatkowski, Cordelia Langford, John Sillitoe on behalf of the Wellcome Sanger Institute COVID-19 Surveillance Team |
| EPI_ISL_674180 | Lighthouse Lab in Milton Keynes | Wellcome Sanger Institute for the COVID-19 Genomics UK (COG-UK) Consortium | The Lighthouse Lab in Milton Keynes and Alex Alderton, Roberto Amato, Sonia Goncalves, Ewan Harrison, David K. Jackson, Ian Johnston, Dominic Kwiatkowski, Cordelia Langford, John Sillitoe on behalf of the Wellcome Sanger Institute COVID-19 Surveillance Team |
| EPI_ISL_683466 | Respiratory Virus Unit, Microbiology Services Colindale, Public Health England | COVID-19 Genomics UK (COG-UK) Consortium | PHE Covid Sequencing Team |
| EPI_ISL_702193, EPI_ISL_719083, EPI_ISL_719450 | Lighthouse Lab in Cambridge | Wellcome Sanger Institute for the COVID-19 Genomics UK (COG-UK) Consortium | Rob Howes, The Lighthouse Lab in Cambridge and Alex Alderton, Roberto Amato, Sonia Goncalves, Ewan Harrison, David K. Jackson, Ian Johnston, Dominic Kwiatkowski, Cordelia Langford, John Sillitoe on behalf of the Wellcome Sanger Institute COVID-19 Surveillance Team |
| EPI_ISL_720147, EPI_ISL_735825 | Lighthouse Lab in Milton Keynes | Wellcome Sanger Institute for the COVID-19 Genomics UK (COG-UK) Consortium | The Lighthouse Lab in Milton Keynes and Alex Alderton, Roberto Amato, Sonia Goncalves, Ewan Harrison, David K. Jackson, Ian Johnston, Dominic Kwiatkowski, Cordelia Langford, John Sillitoe on behalf of the Wellcome Sanger Institute COVID-19 Surveillance Team |
| EPI_ISL_738108 | Instituto Nacional de Saude (INSA) | Instituto Nacional de Saude (INSA) | Borges et al |
| EPI_ISL_756739 | Lighthouse Lab in Milton Keynes | Wellcome Sanger Institute for the COVID-19 Genomics UK (COG-UK) Consortium | The Lighthouse Lab in Milton Keynes and Alex Alderton, Roberto Amato, Sonia Goncalves, Ewan Harrison, David K. Jackson, Ian Johnston, Dominic Kwiatkowski, Cordelia Langford, John Sillitoe on behalf of the Wellcome Sanger Institute COVID-19 Surveillance Team |
| EPI_ISL_760287 | Lighthouse Lab in Glasgow | Wellcome Sanger Institute for the COVID-19 Genomics UK | Harper VanSteenhouse, Yumi Kasai, David Gray, Carol Clugston, Anna Dominiczak and Alex Alderton, Roberto Amato, Sonia Goncalves, Ewan Harrison, |

| | | | |
|---|---|---|---|
| | | (COG-UK) Consortium | David K. Jackson, Ian Johnston, Dominic Kwiatkowski, Cordelia Langford, John Sillitoe on behalf of the Wellcome Sanger Institute COVID-19 Surveillance Team |
| EPI_ISL_783232 | Lighthouse Lab in Milton Keynes | Wellcome Sanger Institute for the COVID-19 Genomics UK (COG-UK) Consortium | The Lighthouse Lab in Milton Keynes and Alex Alderton, Roberto Amato, Sonia Goncalves, Ewan Harrison, David K. Jackson, Ian Johnston, Dominic Kwiatkowski, Cordelia Langford, John Sillitoe on behalf of the Wellcome Sanger Institute COVID-19 Surveillance Team |
| EPI_ISL_783422 | Lighthouse Lab in Cambridge | Wellcome Sanger Institute for the COVID-19 Genomics UK (COG-UK) Consortium | Rob Howes, The Lighthouse Lab in Cambridge and Alex Alderton, Roberto Amato, Sonia Goncalves, Ewan Harrison, David K. Jackson, Ian Johnston, Dominic Kwiatkowski, Cordelia Langford, John Sillitoe on behalf of the Wellcome Sanger Institute COVID-19 Surveillance Team |
| EPI_ISL_799577 | Lighthouse Lab in Milton Keynes | Wellcome Sanger Institute for the COVID-19 Genomics UK (COG-UK) Consortium | The Lighthouse Lab in Milton Keynes and Alex Alderton, Roberto Amato, Sonia Goncalves, Ewan Harrison, David K. Jackson, Ian Johnston, Dominic Kwiatkowski, Cordelia Langford, John Sillitoe on behalf of the Wellcome Sanger Institute COVID-19 Surveillance Team |
| EPI_ISL_802384 | MSHS Clinical Microbiology Laboratories | MSHS Pathogen Surveillance Program | Ana S. Gonzalez-Reiche, Hala Alshammary, Mitchell J. Sullivan, Brianne Ciferri, Ajay Obla, Angela Amoako, Mahmoud Awawda, Elena Hirsch, Ashley S. Salimbangon, Levy Sominsky, Katherine Beach, Kayla Russo, Charles Gleason, Shelcie Fabre, Giulio Kleiner, Zenab Khan, Bremy Alburquerque, Adriana van de Guchte, Komal Srivastava, Matthew M. Hernandez, Jayeeta Dutta, Denise Jurczyszak, Emily Ferreri, Rachel Chernet, Nancy Francoeur, Betsaida Salom Melo, Irina Oussenko, Gintaras Deikus, Juan Soto, Shwetha Hara Sridhar, Ying-Chih Wang, Kathryn Twyman, Andrew Kasarskis, Deena R. Altman, Robert Sebra, Adolfo García-Sastre, Marta Luksza, Gopi Patel, Sarah Schaefer, Melissa Gitman, Michael D. Nowak, Alberto Paniz-Mondolfi, Emilia Mia Sordillo, Viviana Simon, Harm van Bakel |
| EPI_ISL_804972 | Hospital Comarcal de Melilla | Instituto de Salud Carlos III | Iglesias-Caballero, M. Molinero Calamita, M. González-Esguevillas, M. Camarero, S. Pozo, F. Casas, I. Jiménez, P. Jiménez, M. Zaballos, A. Monzón, S. Varona, S. Juliá, M. Cuesta, I, J. López |
| EPI_ISL_822254 | Lighthouse Lab in Alderley Park | Wellcome Sanger Institute for the COVID-19 Genomics UK (COG-UK) Consortium | Jacquelyn Wynn, Mairead Hyland, The Lighthouse Lab in Alderley Park and Alex Alderton, Roberto Amato, Sonia Goncalves, Ewan Harrison, David K. Jackson, Ian Johnston, Dominic Kwiatkowski, Cordelia Langford, John Sillitoe on behalf of the Wellcome Sanger Institute COVID-19 Surveillance Team |
| EPI_ISL_835147 | Lighthouse Lab in Glasgow | Wellcome Sanger Institute for the COVID-19 Genomics UK (COG-UK) Consortium | Harper VanSteenhouse, Yumi Kasai, David Gray, Carol Clugston, Anna Dominiczak and Alex Alderton, Roberto Amato, Sonia Goncalves, Ewan Harrison, David K. Jackson, Ian Johnston, Dominic Kwiatkowski, Cordelia Langford, John Sillitoe on behalf of the Wellcome Sanger Institute COVID-19 Surveillance Team |
| EPI_ISL_835999 | Lighthouse Lab in Milton Keynes | Wellcome Sanger Institute for the COVID-19 Genomics UK (COG-UK) Consortium | The Lighthouse Lab in Milton Keynes and Alex Alderton, Roberto Amato, Sonia Goncalves, Ewan Harrison, David K. Jackson, Ian Johnston, Dominic Kwiatkowski, Cordelia Langford, John Sillitoe on behalf of the Wellcome Sanger Institute COVID-19 Surveillance Team |
| EPI_ISL_838018 | Department of Pathology, University of Cambridge | COVID-19 Genomics UK (COG-UK) Consortium | Aminu S. Jahun, Yasmin Chaudhry, Grant Hall, Iliana Georgana, Myra Hosmillo, Martin D. Curran, Malte Pinckert, Surendra Parmar, Ian Goodfellow |
| EPI_ISL_840744 | Originating lab: Wales Specialist Virology Centre Sequencing lab: Pathogen Genomics Unit | Public Health Wales Microbiology Cardiff Wales Specialist Virology Centre | Catherine Moore, Johnathan Evans, Laura Gifford, Malorie Perry, Simon Cottrell, Angela Marchbank, Alec Birchley, Alexander Adams, Amy Gaskin, Bree Gatica-Wilcox, Jason Coombes, Joel Southgate, Lauren Gilbert, Lee Graham, Nicole Pacchiarini, Sara Kumziene-Summerhayes, Sarah Taylor, Sophie Jones, Sara Rey, Matthew Bull, Joanne Watkins, Sally Corden, Tom Connor |
| EPI_ISL_842652 | Laboratorio de Biología Molecular Hospital Pedro de Elizalde | Grupo de Genómica y Bioinformática del Instituto de Investigación de la Cadena Láctea CONICET-INTA on behalf of 'Proyecto Argentino Interinstitucional de genomica de SARS-CoV-2' (PAIS Consortium) | Amadio, AF, Eberhardt, MF; Irazoqui, M; Indart, J; Rocovich, J; Montoto Piazza, L; Wenk, G; Martín, ME; Sanchez, MF; Marchetti, P; Morandi, F; Sueiro, ML; Claps, A; Bressan, L; Torres, FJ; Chamorro, J; Gondolessi, J; Gómez, ML; Diaz, B; Rosales, D; Alegre, F; Zamora, N; Osaba, E; Paez, E; Lorenzo, F; Torres, C; Aulicino, P; König, G; Alexay, S; Natale, M; Valinotto, L; Lusso, S; Goya, S; Nabaes Jodar, MS; Viegas, M. |
| EPI_ISL_843045, EPI_ISL_843060 | Barts Health NHS Trust | COVID-19 Genomics UK (COG-UK) Consortium | CUTINO-MOGUEL, Maria-Teresa; HARRINGTON, David; OWOYEMI, Dola; SHYLINI, Raghavendran; BROAD, Claire; KELE, Beatrix |
| EPI_ISL_863793 | Lighthouse Lab in Milton Keynes | Wellcome Sanger Institute for the COVID-19 Genomics UK (COG-UK) Consortium | The Lighthouse Lab in Milton Keynes and Alex Alderton, Roberto Amato, Sonia Goncalves, Ewan Harrison, David K. Jackson, Ian Johnston, Dominic Kwiatkowski, Cordelia Langford, John Sillitoe on behalf of the Wellcome Sanger Institute COVID-19 Surveillance Team |
| EPI_ISL_924084 | Virology Department, Sheffield Teaching Hospitals NHS Foundation Trust/Department of Infection, Immunity and Cardiovascular Disease, The Medical School, University of Sheffield | COVID-19 Genomics UK (COG-UK) Consortium | Thushan de Silva, Matthew Parker, Nikki Smith, Adri Angyal, Rebecca Brown, Luke Green, Rachel Tucker, Paul Parsons, Danielle Groves, Katie Johnson, Laura Carrilero, Alex Keeley, Dave Partridge, Matthew Wyles, Benjamin Lindsey, Mehmet Yavuz, Mohammad Raza, Cariad Evans |
| EPI_ISL_977235 | ULSS 5 Polesana | Istituto Zooprofilattico Sperimentale delle Venezie | Adelaide Milani, Alessia Schivo, Annalisa Salviato, Erika Giorgia Quaranta, Ambra Pastori, Bianca Zecchin, Alice Fusaro, Isabella Monne, Calogero Terregino, Antonia Ricci |
| EPI_ISL_981967 | Microbiology Service, Hospital Universitario Clinico San Cecilio, Granada | Microbiology Service, Hospital Universitario Clinico San Cecilio, Granada | Adolfo de Salazar, Natalia Chueca, Laura Viñuela, Ana Fuentes, Federico García |
| EPI_ISL_997769 | University College London, Great Ormond Street Hospital for Children NHS Foundation Trust, Imperial College Healthcare NHS Trust | COVID-19 Genomics UK (COG-UK) Consortium | Sergi Castellano, Rachel Williams, Mark Kristiansen, Paola Resende Silva, Sunando Roy, Tony Brooks, Helena Tutill, Paola Niola, Patricia Dyal, Charlotte Williams, Leysa Forrest, Yasmin Panchbhaya, Jacqueline Findlay, Samuel Weeks, Julianne Brown, Kathryn Harris, Paul Randell, James Price, Alison Holmes, Judith Breuer |
